# Changes in cancer prevention and management and patient needs during the COVID-19 pandemic: An umbrella review of systematic reviews

**DOI:** 10.1101/2022.12.18.22283642

**Authors:** Taulant Muka, Joshua J X Li, Sahar J. Farahani, John P.A. Ioannidis

## Abstract

**Introduction:** The COVID-19 pandemic led to relocation and reconstruction of health care resources and systems, and to a decrease in healthcare utilization, and this may have affected the treatment, diagnosis, prognosis, and psychosocial well-being of cancer patients.

**Objective:** To summarize and quantify the evidence on the impact of the COVID-19 pandemic on the full spectrum of cancer care.

**Methods:** We performed an umbrella review to summarize and quantify the findings from systematic reviews on impact of the COVID-19 pandemic on cancer treatment modification, delays, and cancellations; delays or cancellations in screening and diagnosis; psychosocial well-being, financial distress, and use of telemedicine as well as on other aspects of cancer care. PubMed was searched for relevant systematic reviews with or without meta-analysis published before November 29^th^, 2022. Abstract, full text screening and data extraction were performed by two independent reviewers. AMSTAR-2 was used for critical appraisal of included systematic reviews.

**Results:** 45 systematic reviews evaluating different aspects of cancer care were included in our analysis. Most reviews were based on observational studies judged to be at medium and high risk of bias. Only 2 of the included reviews had high or moderate scores based on AMSTAR-2. Findings suggest treatment modifications in cancer care during the pandemic versus the pre-pandemic period were based on low level of evidence. Different degrees of delays and cancellations in cancer treatment, screening and diagnosis were observed, with low-and-middle income countries and countries that implemented lockdowns being disproportionally affected. A shift from in-person appointments to telemedicine use was observed, but utility of telemedicine, challenges in implementation and cost-effectiveness in different areas of cancer care were little explored. Evidence was consistent in suggesting psychosocial well-being (e.g., depression, anxiety, and social activities) of cancer patients deteriorated, and cancer patients experienced financial distress, albeit results were in general not compared to pre-pandemic levels. Impact of cancer care disruption during the pandemic on cancer prognosis was little explored.

**Conclusion:** Substantial but heterogenous impact of COVID-19 pandemic on cancer care has been observed. Evidence gaps exist on this topic, with mid- and long-term impact on cancer care being most uncertain.

## INTRODUCTION

The coronavirus disease 2019 (COVID-19) pandemic and the mitigation measures that were undertaken posed major challenges to cancer care. The rapid spread of COVID-19 and early data showing patients with cancer were at increased risk of morbidity and mortality after Severe Acute Respiratory Syndrome Coronavirus 2 (SARS-CoV-2) infection, prompted changes in healthcare delivery^1^. These changes included reduction of medical activities, reallocation of healthcare workers, shifting in-person appointments to remote consultations, and limiting access of patients to care facilities^2^.

Concerns have been raised that disruption of health care services might have had multidimensional impact in cancer care. Indeed, several studies have described delays and cancellation in treatment, screening, and diagnosis^3-5^. For example, two meta-analyses showed that during the pandemic there was a ∼50% reduction in breast and cervical cancer screening, and that there was 18.7% reduction for all cancer treatments, with surgical treatment showing the highest reduction^3 4^. In addition, several studies have highlighted deterioration of psychological well-being of cancer patients, and psychological, ethical, spiritual, and financial needs of cancer patients were also affected^6 7^.

Several systematic reviews have examined the impact on cancer care, but they evaluated different outcomes and periods of the pandemic^3 4 8-14^. Thus, it would be essential to put together these systematic reviews, particularly assessing the methodological rigor of the evidence and summarizing systematically the main findings in terms of the magnitude of the impact and the uncertainty thereof. To achieve these goals, we performed an umbrella review of systematic reviews.

## METHODS

We performed an umbrella review following the recent published guideline^15^, and for reporting we adhered to the Preferred Reporting Items for Overviews of Reviews-PRIOR checklist^16^ (**Appendix 1**). The protocol has been registered with the Open Science Framework (https://osf.io/qjgxv)

### Search Strategy

Literature search was performed in PubMed using the search strategy in **Appendix 2**. No language restriction was applied. We searched for studies published until November 3^rd^, 2022; an update of the search was performed until November 29^th^, 2022. References cited in the final included studies for analysis were further screened to identify other relevant publications.

### Screening, Study selection and Eligibility criteria

Retrieved items were first screened based on the title and abstract and potentially eligible references were then screened in full text. Screening was performed by two reviewers and in case of discrepancies, a final decision to include or exclude was settled with discussion. We included studies if they fulfilled all the following criteria: (i) were systematic reviews with our without meta-analysis or individual participant meta-analysis; (ii) included individuals diagnosed with any type of cancer and at any cancer stages (early to advanced), or individuals targeted for cancer screening; (iii) assessed the impact of the COVID-19 pandemic, and thus had data collected during the pandemic period (2020-2022) (the included studies may nevertheless have used also control pre-pandemic periods in order to assess the magnitude of change during the pandemic); and assessed any of the following outcomes: delay/cancellation of treatment (overall, and per specific treatment); modification of treatment (overall, and per specific treatment); delayed/cancelled screening (overall and per specific type of screening); reduced diagnoses (overall and per specific diagnosis); reduced uptake of HPV vaccination; psychological needs; ethical needs; social needs; financial burden and distress; social impact/ isolation; psychological distress; use of telehealth/virtual visits; tobacco use and cessation and other aspects of cancer care such as impact of the COVID-19 pandemic on prognosis.

### Data extraction and Critical appraisal

The data extraction was performed by one of the authors and the extracted data were further checked by two other authors; differences were settled by discussion. In case an eligible article included data from several diseases, when feasible, we extracted information only on cancer-related outcomes of our interest. For each eligible systematic review we extracted the following information: authors, year of publication, type of studies considered (design), number of eligible studies, COVID-19 period covered (until when), whether it has considered studies with pre-pandemic controls (yes exclusively/yes for some/not at all), the location(s) that were eligible (countries/areas/global), the outcomes examined and for which cancers each outcome was examined, and methods of analysis and heterogeneity (if provided).

For each systematic review we extracted information on whether the authors used any previously validated tool or any other set of extracted items to assess the methodological rigor of the included studies. If yes, we recorded the tool used and the main conclusions of the assessment were grouped in the broad categories: most studies were weak in methodological rigor, most studies were strong in methodological rigor, or mixed/ intermediate pattern between the other two categories. Two reviewers assessed methodological rigor of the included systematic reviews using AMSTAR-2 tool^17^; any discrepancies were settled with the help of a third reviewer. AMSTAR-2 is based on a 16 item or domain checklist, with seven of these items considered critical the overall validity of a review. The domains considered critical are: (i) protocol registration before starting the review; (ii) adequate and comprehensive search of the literature; (iii) providing justification for the exclusion of individual studies; (iv) risk of bias assessment of the studies included in the review; (v) use of appropriate statistical methods in performing a meta-analysis; (vi) accounting for risk of bias when interpreting the results; (vii) and evaluation of the presence and impact of publication bias.

### Statistical analysis

Due to high heterogeneity in the designs, study questions, outcomes, and metrics, a descriptive analysis was performed. Separate tables were created for the methodological appraisal of the systematic reviews, the methodological appraisal of the studies in each systematic review, for the characteristics and subject matter information of each systematic review, and for the final conclusions of each systematic review. Limitations and areas of limited evidence were noted.

## RESULTS

Our search strategy identified 649 citations. Based on title and abstract screening, we retrieved full texts of 74 articles for further screening. Of those, 29 articles did not meet our eligibility criteria, thus leaving 45 articles to be included in our final analysis. **Figure 1** summarizes our screening procedure. No additional study was found from screening of references of the included studies.

**Figure 1.**
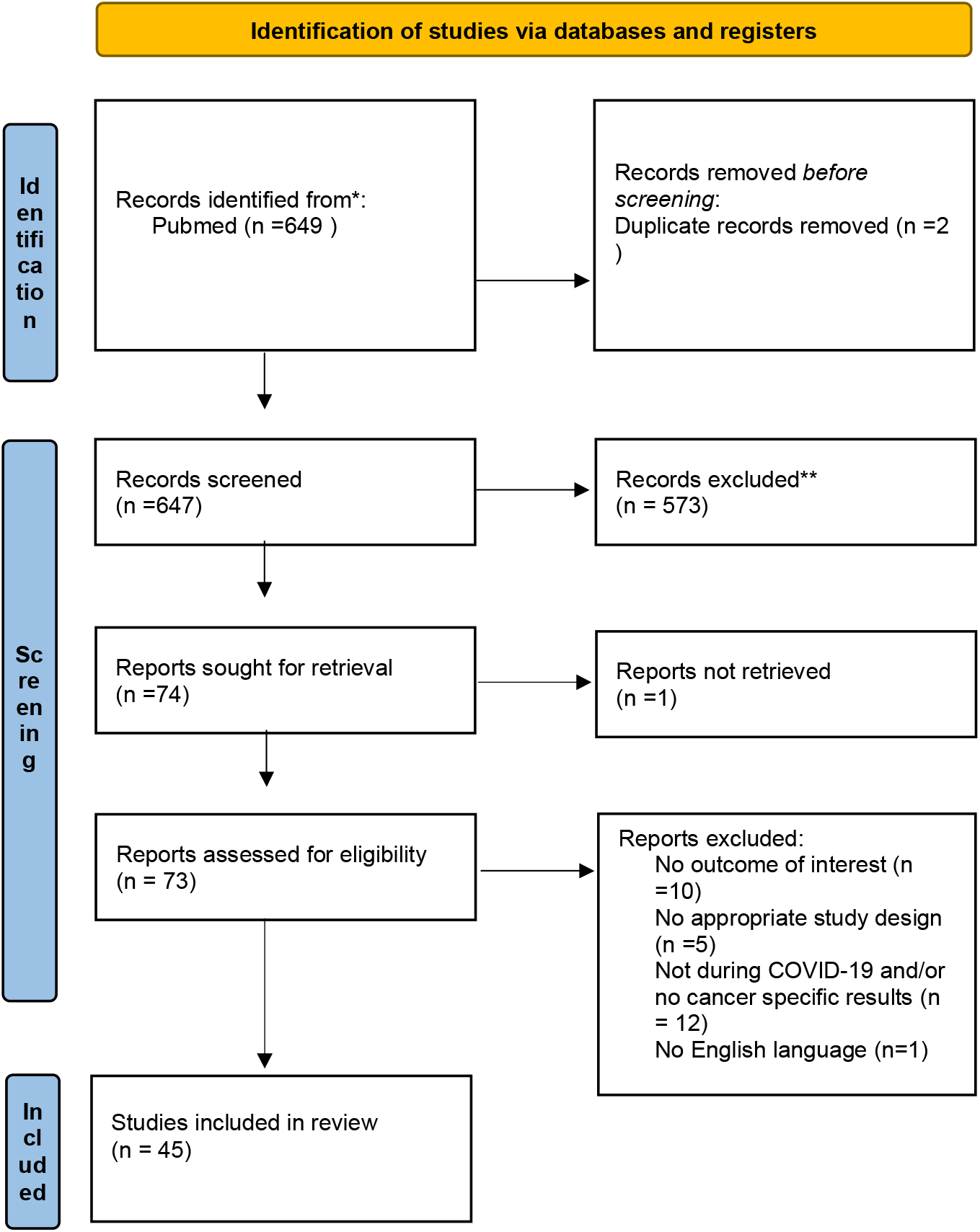
Flowchart of Identification, Screening, Eligibility, Inclusion, and Exclusion of Retrieved Studies

### Characteristics of the included systematic reviews

Of the 45 included systematic reviews, 13 articles also included a quantitative analysis/meta-analysis with one being individual participant meta-analysis.^2-14 18-49^ Other key characteristics of the 45 systematic reviews are shown in **Table 1** (more extensive details appear in **Appendix 3 and Appendix 4**). The median number of bibliographic databases/data sources that were searched was 3.5; the most searched databases were PubMed (n=23), Medline (n=22), Embase (n=22), Scopus (n=17), Web of Science (n=13) and The Cumulative Index to Nursing and Allied Health Literature-CINAHL database (n=10). One review searched for mobile applications using the iOS App Store and Android Google Play^23^. The median number of studies included in the systematic reviews was 31 (interquartile range, 15.5; 49). Most reviews provided data from different countries, while only two studies focused on data from India^29^ and Italy^35^ exclusively. The type of study designs included across reviews varied, but most reviews included data from observational study designs of cross-sectional and retrospective nature. Twenty reviews focused/reported exclusively on studies that include pre-pandemic controls.

**Table 1.** Characteristics of included systematic reviews

| Author, year of publication | Meta-analysis | Number of included studies | Countries | Pre-pandemic controls | Cancer types | Aspects assessed | Last search |
| --- | --- | --- | --- | --- | --- | --- | --- |
| Adham, 2022<sup>18</sup> | No | 5 | Globally | No | H&N | MT, O | 15-Jul-20 |
| Alom, 2021<sup>19</sup> | No | 72 | Globally | No | All | MT, TL, O | 1-Sep-20 |
| Ayubi, 2021<sup>20</sup> | Yes | 34 | Globally | No | All | PSND, O | 3-Jan-21 |
| Dhada, 2021<sup>2</sup> | No | 19 | IT, US, UK, NL | No | ALL | DCT, DCS, PSND, TL, FBD, SIA | 1-Dec-20 |
| Donkor, 2021<sup>8</sup> | No | 11 | CN, IR, BR, ZA | No | ALL | O | 3-Aug-20 |
| Garg, 2020<sup>21</sup> | No | 212 | Globally | No | ALL | MT | 2-May-20 |
| Gascon, 2020<sup>9</sup> | No | 23 | Globally | No | H&N | MT, O | 1-May-20 |
| Hojaij, 2020<sup>10</sup> | No | 35 | Globally | No | H&N,OT<br>O | MT, TL, O | 31-Dec-20 |
| Jammu, 2021<sup>22</sup> | No | 19 | Globally | No | ALL | DCT, PSND, FBD | 27-Aug-20 |
| Kirby, 2022<sup>7</sup> | No | 56 | Globally | No | ALL | PSND, FBD, SIA | 31-Mar-21 |
| Legge, 2022<sup>11</sup> | No | 18 | Globally | No | ALL | PSND, FBD, SIA | 25-May-22 |
| Lu, 2021<sup>23</sup> | No | 41* | NA | No | ALL | TL | 1-May-20 |
| Moemenimovahed, 2021<sup>24</sup> | No | 55 | Globally | No | ALL | PSND | 30-Jun-21 |
| Mostafaei, 2022<sup>25</sup> | No | 22 | Globally | No | ALL | TL | 1-Jun-21 |
| Moujaess, 2020<sup>26</sup> | No | 88 | Globally | No | ALL | DCT, O | 15-Apr- |
|  |  |  |  |  |  |  | 20 |
| Muls, 2022<sup>27</sup> | No | 51 | Globally | No | ALL | PSND | 1-Oct-21 |
| Murphy A, 2022<sup>12</sup> | No | 37 | Globally | No | ALL | TL | 31-Mar-21 |
| Pacheco, 2021<sup>28</sup> | No | 9 | US, IT, CN, SP, UK, IR | No | ALL | DCT, O | NP |
| Rohilla, 2021<sup>29</sup> | No | 6 | IN | No | ALL | PSND, O | 3-Feb-21 |
| Salehi, 2022<sup>30</sup> | No | 16 | Globally | No | ALL | TL | 1-Apr-21 |
| Sun P, 2021<sup>31</sup> | No | 6 | IT, AM, UK | No | BC | MT | 1-Feb-21 |
| Zapala, 2022<sup>32</sup> | No | 160 | NP | No | ALL | DCT, PSND, TL | NP |
| Zhang, 2022<sup>6</sup> | Yes | 40 | Globally | No | ALL | PSND | 31-Jan-22 |
| Alkatoul, 2021<sup>33</sup> | No | 16 | US, TW, BE, NL, JP, IT, UK, AS, CA | Yes | ALL | DCS, RD | 28-Dec-20 |
| Di Cosimo, 2022<sup>34</sup> | Yes | 56 | Globally | Yes | ALL | MT, DCT, TL, O | 11-Dec-20 |
| Fancellu, 2022<sup>35</sup> | No | 7 | IT | Yes | CRC | DCS, RD | 31-Jan-22 |
| Ferrar, 2022<sup>36</sup> | No | 33 | Globally | Yes | CV | DCT, DCS, RD, RHPV | 8-Feb-22 |
| Gadsden, 2022<sup>13</sup> | No | 17 | IN, SL, BA | Yes | ALL | DCT, O | 15-Dec-21 |
| Hesary, 2022<sup>37</sup> | No | 22 | IT, UK, PG, NL, CN, IN, JP, TU, IR, SN | Yes | GA | MT, DCS, RD, PSND | 31-Dec-21 |
| Lignou, 2022<sup>38</sup> | No | 32 | Globally | Yes | PC | DCT, RD, TL | 1-Aug-21 |
| Majeed, 2021<sup>14</sup> | No | 60 | Globally | Yes, but NS | PC | DCT, RD, TL | 3-Nov-21 |
| Mayo, 2021<sup>39</sup> | Yes | 13 | IT, AU, TW, US, FR, NL | Yes | ALL | DCT, DCS | 10-Feb-21 |
| Mazidimoradi, 2021<sup>40</sup> | No | 43 | Globally | Yes | CRC | MT, DCT, RD | 1-Jun-21 |
| Mazidimoradi, 2022<sup>41</sup> | No | 25 | Globally | Yes | CRC | DCS | 1-Jun-21 |
| Ng, 2022<sup>42</sup> | Yes | 31 | Globally | Yes | BC | DCS, RD | 1-Oct-20 |
| Nikolopoulos, 2022<sup>5</sup> | No | 15 | Globally | Yes, but NS | GC | MT, DCT, RD, PSND | 10-Feb-21 |
| Pararas, 2022<sup>43</sup> | Yes | 10 | Globally | Yes | CRC | O | NP |
| Riera, 2021<sup>44</sup> | No | 62 | Globally | Yes | ALL | DCT | NP |
| Sarich, 2021<sup>45</sup> | Yes | 44 | Globally | Yes | NA | RF | 5-Nov-20 |
| Sasidharanpillai, 2022<sup>46</sup> | Yes | 7 | SL, IT, CA, SC, BE, US | Yes | CV | DCT, RD | 1-Sep-21 |
| Tang, 2022<sup>47</sup> | Yes | 14 | TU, CN, UK, IT, DN, AS, AU | Yes | CRC | O | 12-Jan-22 |
| Teglia, 2022<sup>3</sup> | Yes | 39 | Globally | Yes | BC, CRC, CV | DCT, RD | 12-Dec-21 |
| Teglia, 2022<sup>4</sup> | Yes | 47 | Globally | Yes | ALL | DCT | 12-Dec-21 |
| Thomson, 2020<sup>48</sup> | Yes | 54 | NP | Yes | ALL | O | 1-Jun-21 |
| Vigliar, 2020<sup>49</sup> | Yes | 41** | Globally | Yes | ALL | DCS, RD | 30-Apr-20 |

AM, America; BC, AS, Austria; AU, Australia; BA, Bangladesh; BC, breast cancer; BE, Belgium; BR, Brazil; CA, all cancers or Canada; CN, China; CRC, colorectal cancer; CV, cervical cancer; DN, Denmark; FR, France; GA, gastric cancer; GC, gynecological cancer; H&N, head and neck cancer; IN, India; IR, Iran; IT, Italy; JP, Japan; NA, not applicable; NL, Netherlands; NP, not provided; OTO, otorhinolaryngology cancer; PC, pediatric cancer; PG, Portugal; SC, Scotland; SL, Slovenia or Sri Lanka; SN, Singapore; SP, Spain; TU, Turkey; TW, Taiwan; UK, United Kingdom; United States; ZA, Zambia;
\*apps; \*\*respondents

Seventeen reviews provided data only on site-specific cancers, while the rest for any cancer-site with or without data on site-specific cancers. Seventeen reviews assessed only one aspect of cancer care, while the rest examined two or more of our pre-defined outcomes. The date of last search varied from April 2020 to May 2022, with 15 reviews ending searches during 2020, 21 during 2021 and 5 during 2022; 4 reviews did not provide information on date of last search.

### Risk of bias of primary studies included in the systematic reviews and GRADE assessments

Of the 45 reviews, 31 assessed risk of bias of the included studies (**Table 2** and details in **Appendix 5**). Thirteen different risks of bias checklists were used, and the most common checklists used to assess methodological rigor were Newcastle-Ottawa Scale (NOS) (n=10) and Joanna Briggs Institute tools (n=7). Of the systematic reviews that assess methodological rigor of the individual studies, 7 concluded strong evidence, 19 mixed evidence, 3 weak evidence and 2 did not provide any results. Excluding the NOS assessments (since NOS has been criticized to not provide accurate assessment of methodological rigor^50^), the respective numbers were 2, 14, 3, and 2. Only two reviews used GRADE (Grading of Recommendations, Assessment, Development and Evaluations), concluding low to moderate certainty in the results.

**Table 2:**
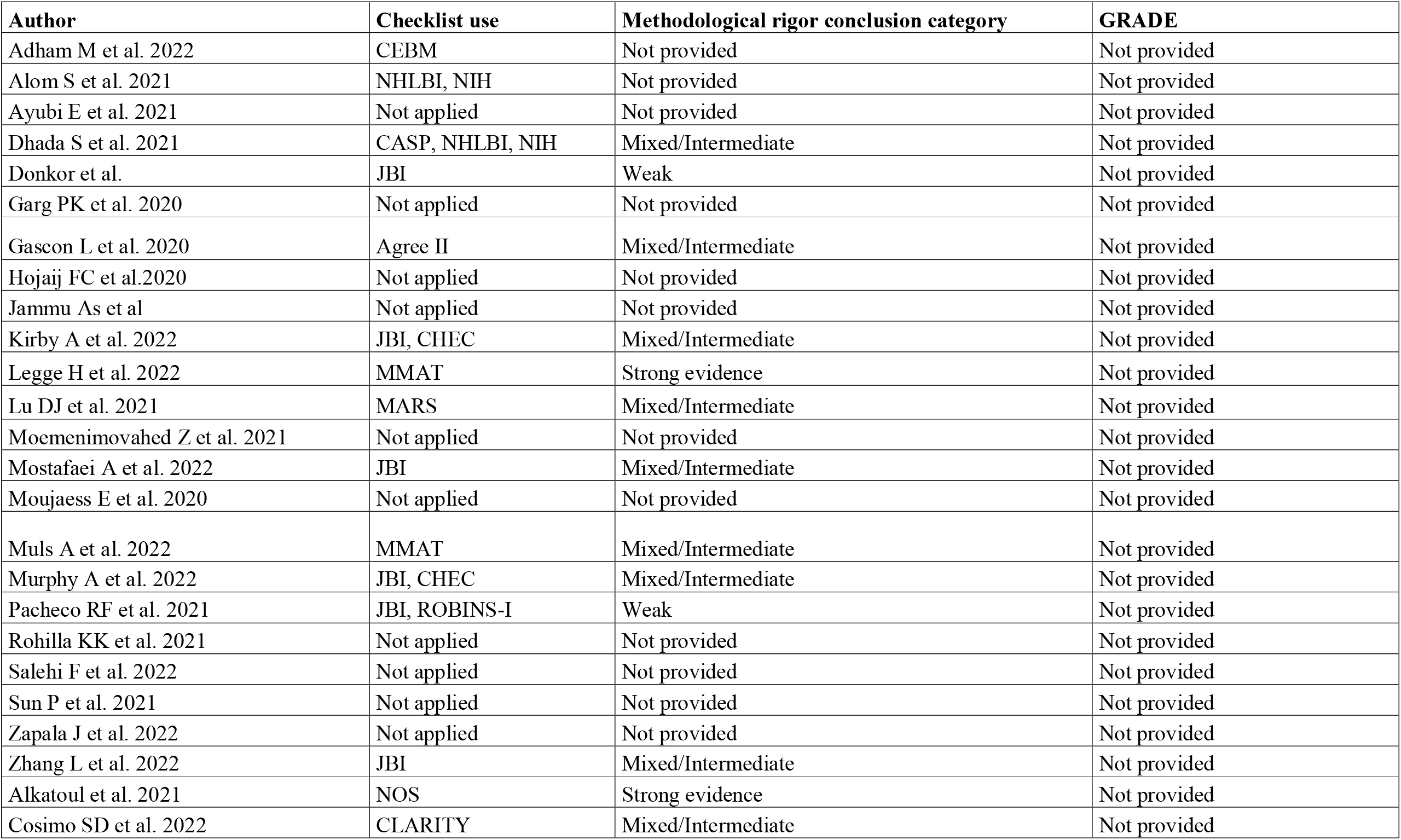
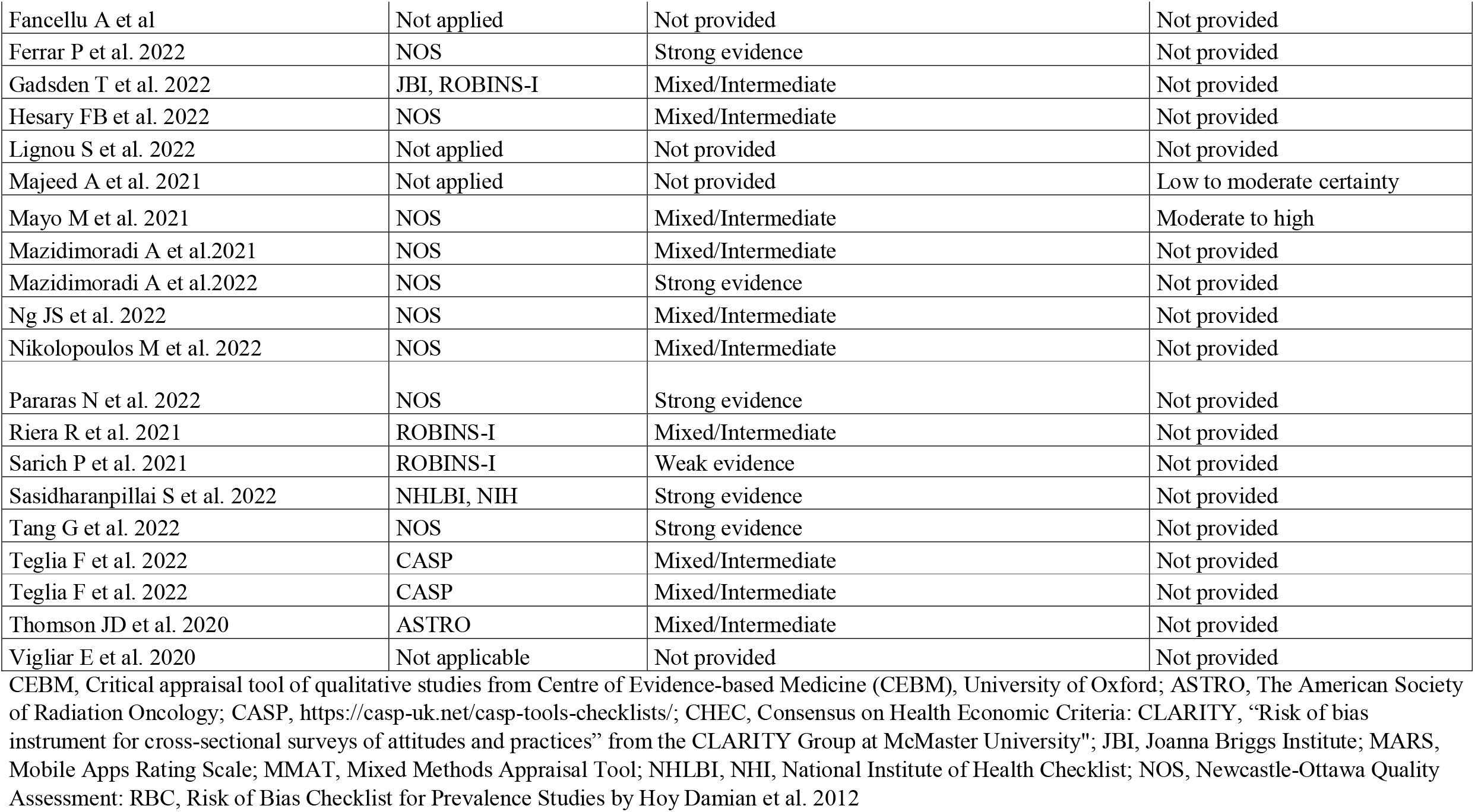
Methodological rigor of included reviews

### Methodological rigor of included systematic reviews

**Table 3** shows the AMSTAR-2 evaluations for the included systematic reviews. Only two reviews scored moderate to high quality, while the rest were evaluated as low or critically low quality due to not meeting one or more of the seven domains considered critical. Most of the studies did not provide the list of excluded studies during the full text screening, and did not account for methodological rigor of included studies when interpreting/discussing the results of the reviews.

**Table 3.** Methodological assessment of the included reviews-AMSTAR 2 evaluation (16 questions)*

| Authors, year of publication | q1 | q2 | q3 | q4 | q5 | q6 | q7 | q8 | q9** | q10 | q11 | q12 | q13 | q14 | q15 | q16 | Overall Assessment |
| --- | --- | --- | --- | --- | --- | --- | --- | --- | --- | --- | --- | --- | --- | --- | --- | --- | --- |
| Adham M et al. 2022 | n | n | n | py | n | n | n | n | y | n | na | na | na | n | na | n | Critical low |
| Alom S et al., 2021 | n | n | n | py | n | y | n | py | y | n | na | na | y | n | na | y | Critical Low |
| Ayubi E et al. 2021 | y | n | n | py | n | n | n | y | n | n | y | n | n | n | y | y | Critical low |
| Dhada S et al. 2021 | n | py | n | py | n | n | n | y | y | n | na | na | n | n | na | y | Critical Low |
| Donkor et al. 2021 | n | n | n | py | y | y | n | y | y | n | na | na | na | n | na | y | Critical low |
| Garg PK et al. 2020 | n | n | n | py | y | y | n | n | n | n | na | na | n | y | na | y | Critical low |
| Gascon L et al. 2020 | y | y | n | y | y | y | n | na | y | y | na | na | na | n | na | y | Low |
| Hojaij FC et al. 2020 | n | n | n | n | n | n | n | n | n | n | na | na | na | n | na | y | Critical low |
| Jammu AS et al. 2021 | n | n | n | py | y | y | n | n | n | n | na | na | n | n | na | y | Critical low |
| Kirby A et al. 2022 | y | py | n | y | n | y | n | py | y | n | na | na | n | n | na | y | Critical Low |
| Legge H et al. 2022 | y | py | y | py | y | y | n | y | y | n | na | na | n | n | na | y | Critical Low |
| Lu DJ et al. 2021 | y | n | na | py | n | n | n | y | na | n | na | na | na | n | na | y | Critical Low |
| Momenimovahed Z et al. 2021 | n | n | n | py | n | n | n | n | n | n | na | na | n | n | na | y | Critical low |
| Mostafaei A et al. 2022 | n | py | n | n | n | n | y | py | y | n | na | na | n | n | na | y | Critical low |
| Muls A et al. 2022 | y | py | y | py | n | y | n | y | y | n | na | na | n | n | na | y | Critical Low |
| Murphy A et al. 2022 | n | n | n | y | n | n | n | y | y | n | na | na | n | n | na | y | Critical low |
| Pacheco RF et al. 2021 | y | y | y | py | y | y | y | py | y | y | na | na | y | n | na | y | High quality |
| Rohilla KK et al. 2021 | n | n | n | py | n | y | n | n | n | n | na | na | n | n | na | y | Critical low |
| Salehi F et al. 2022 | n | n | n | py | y | n | n | n | n | n | na | na | n | n | na | y | Critical low |
| Sun P et al. 2021 | n | n | n | py | n | n | n | n | n | n | na | na | na | n | na | n | Critical low |
| Zapala J et al. 2022 | n | n | n | n | n | n | n | n | n | n | na | na | n | n | na | y | Critical low |
| Zhang L et al. 2022 | y | y | y | py | n | y | n | py | y | n | y | y | y | y | y | y | Low |
| AlkatouI et al. 2021 | n | py | y | py | n | n | n | py | y | n | na | na | n | n | na | y | Critical Low |
| Di Cosimo et al. 2022 | n | n | n | py | y | n | n | y | y | n | y | y | y | y | y | y | Critical low |
| Fancellu A et al. 2022 | y | n | n | n | n | n | n | n | n | n | na | na | n | n | n | n | Critical low |
| Ferrara P et al. 2022 | n | py | n | py | y | y | n | n | y | n | na | na | y | n | na | y | Low |
| Gadsden T et al. 2022 | y | py | n | py | y | n | n | y | y | n | na | na | y | n | na | y | Low |
| Hesary FB et al. 2022 | n | py | n | py | n | n | n | n | y | n | na | na | n | n | na | y | Critical Low |
| Lignou S et al. 2022 | y | n | n | n | y | y | n | y | n | n | na | na | n | n | na | y | Critical low |
| Majeed A et al. 2022 | n | y | n | py | n | y | n | n | py | n | na | na | n | n | na | y | Critical Low |
| Mayo M et al. 2021 | n | y | n | py | y | y | n | n | py | n | n | y | y | n | n | y | Critical low |
| Mazidimoradi A et al.2021 | n | py | n | py | n | n | n | y | y | n | na | na | n | n | na | y | Critical Low |
| Mazidimoradi A et al. 2022 | n | py | n | py | n | n | n | py | y | n | na | na | n | n | na | y | Critical Low |
| Ng JS et al. 2022 | n | py | n | py | n | n | n | py | y | n | y | n | y | y | y | y | Low |
| Nikolopoulos M et al. 2022 | n | py | n | py | n | n | n | n | y | n | na | na | n | n | na | y | Critical Low |
| Pararas N et al. 2022 | n | y | n | y | y | n | n | n | y | n | n | n | n | y | y | y | Critical low |
| Riera R et al. 2021 | n | py | y | py | y | y | y | y | y | y | na | na | n | y | na | y | Moderate quality |
| Sarich P et al. 2022 | y | y | y | py | y | y | n | y | y | n | y | y | n | y | n | y | Critical low |
| Sasidharanpillai et al. 2022 | n | py | n | py | n | n | n | y | y | n | y | y | y | y | y | y | Low |
| Tang G et al. 2022 | y | n | n | n | n | n | n | n | y | py | n | n | n | y | n | y | Critical low |
| Teglia F et al. 2022 | y | py | y | py | y | y | n | n | y | n | n | n | n | n | y | y | Critical low |
| Teglia F et al. 2022 | y | py | y | py | y | y | n | py | y | n | n | n | n | y | n | y | Critical low |
| Thomson JD et al. 2020 | n | n | n | n | n | n | n | n | y | n | y | n | n | n | na | y | Critical low |
| Vigliar E et al., 2020** | na | na | na | na | na | na | na | na | na | na | na | na | na | na | na | na | NA |

n, no; NA, not applicable; py, partially yes; y, yes
\*The review scored yes if study used a checklist to evaluate methodological rigor, and partial yes if only GRADE assessment was provided without applying a checklist for assessing methodological rigor. \*Individual participant meta-analysis and thus not applicable the AMSTAR evaluation
AMSTAR-2 overall assessment rating: high—the review provides an accurate and comprehensive summary of the results of the available studies that addresses the question of interest; moderate—the review has more than one weakness, but no critical flaws. It may provide an accurate summary of the results of the available studies; low—the review has a critical flaw and may not provide an accurate and comprehensive summary of the available studies that address the question of interest; or critically low—the review has more than one critical flaw and should not be relied on to provide an accurate and comprehensive summary of the available studies

### Results and conclusions of systematic reviews and of meta-analyses

The main results and conclusions of the eligible systematic reviews are presented in **Appendices 6-12** for various aspects of cancer care. **Table 4** lists the effect sizes and confidence intervals for the systematic reviews that used formal meta-analysis as well as heterogeneity metrics. **Figure 2** provides a summary of main findings of this umbrella review. Here, we present some key findings for each type of outcome:

**Table 4:**
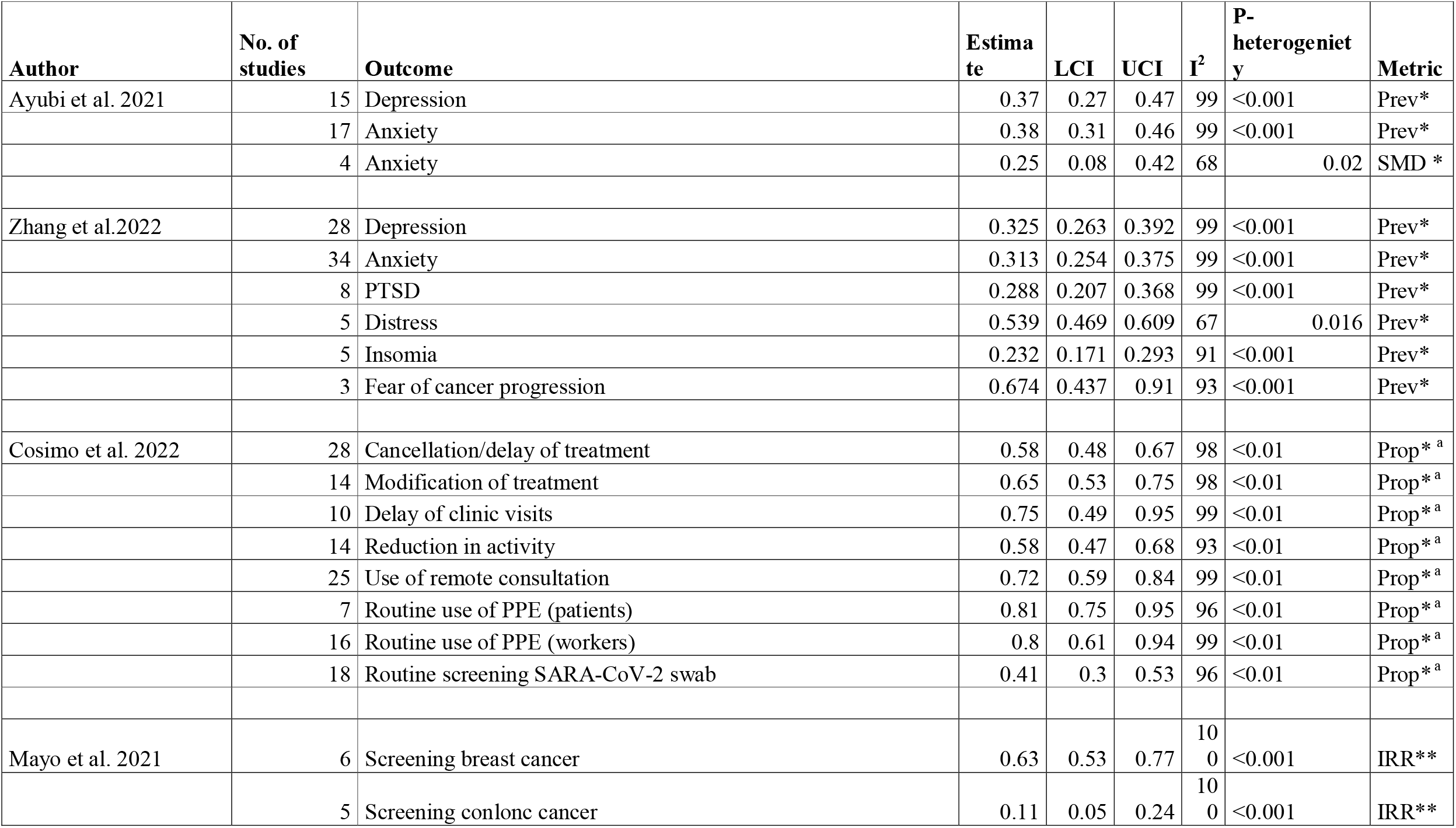
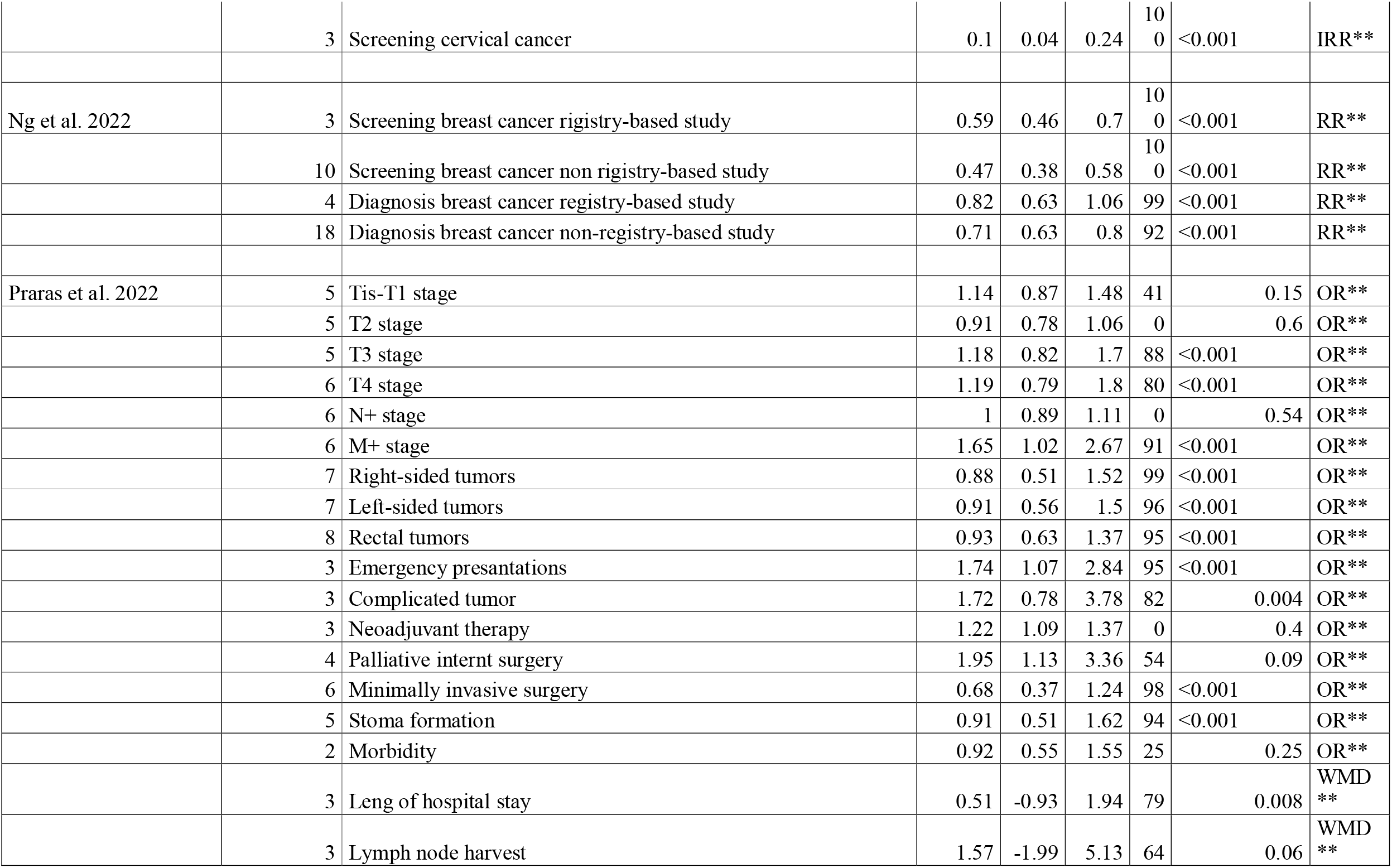
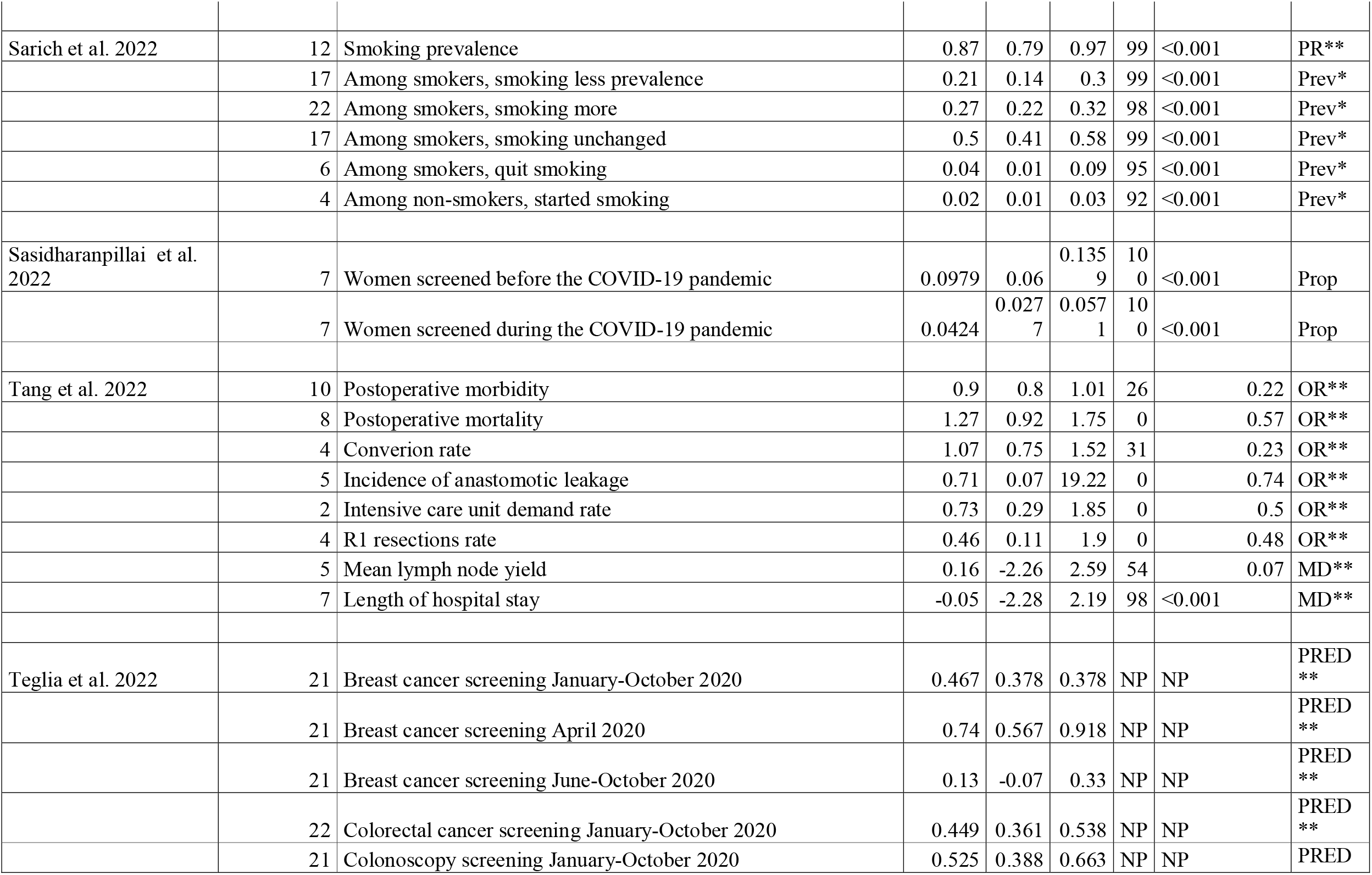
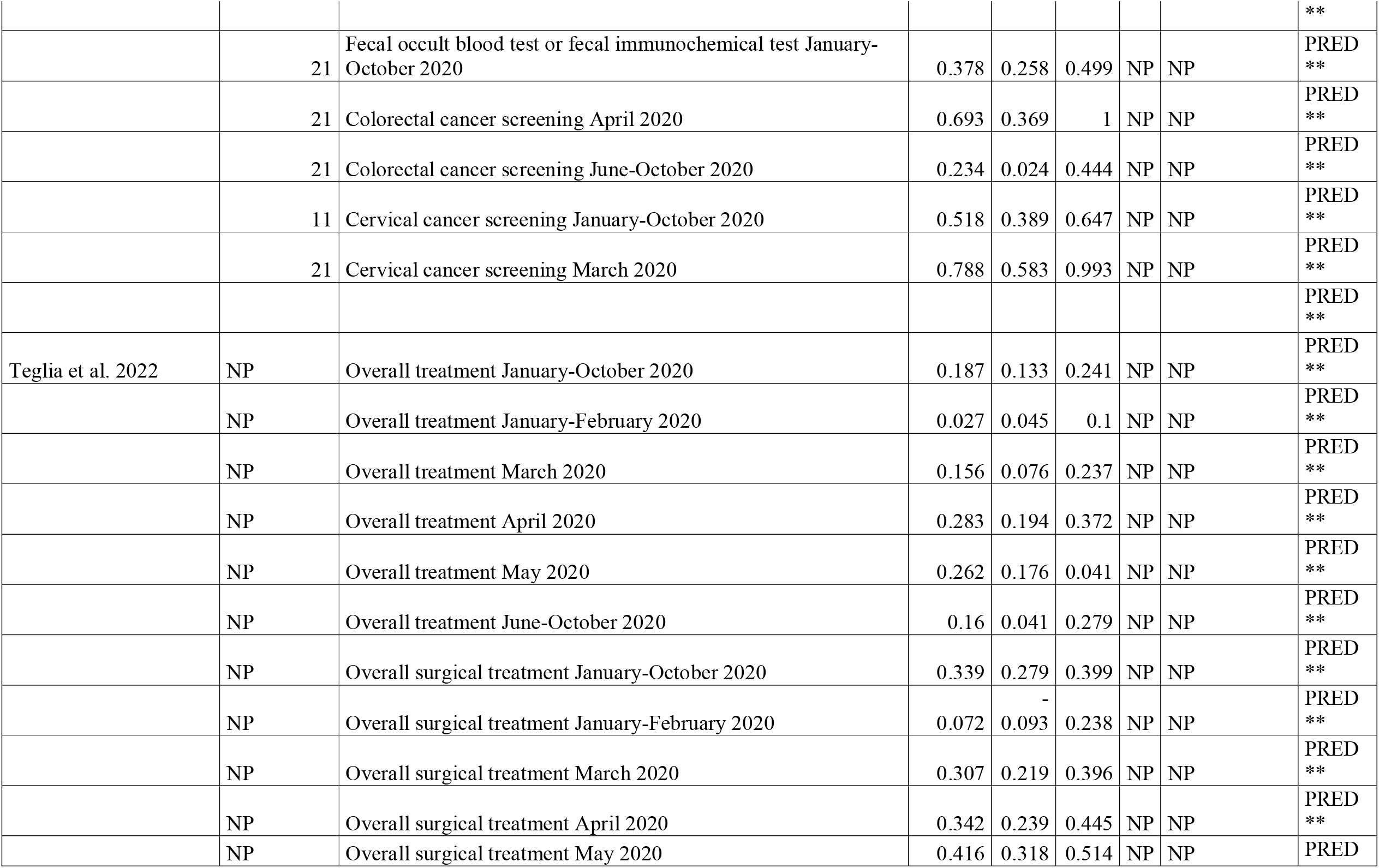
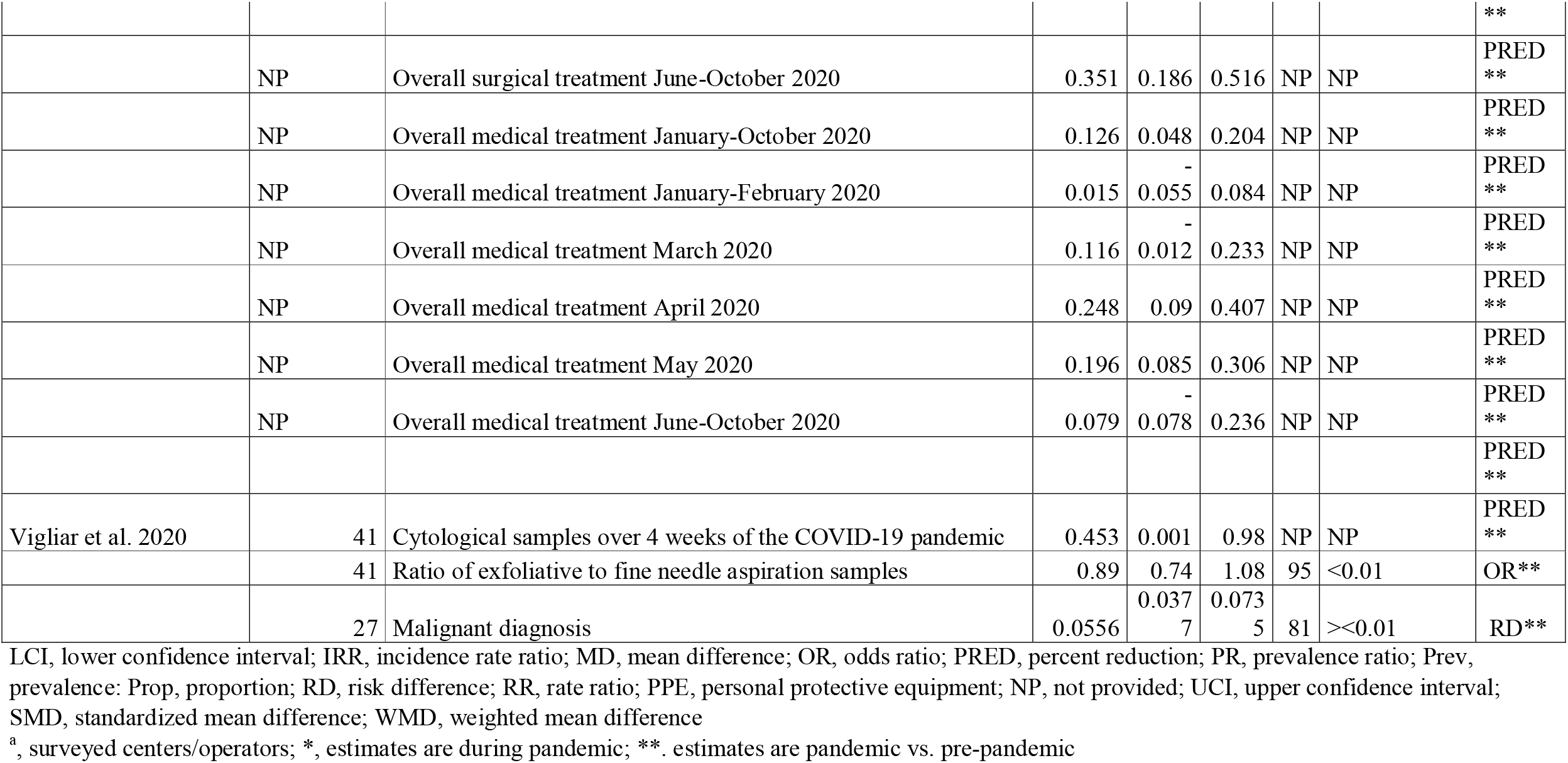
Summary estimates of the meta-analysis included

**Figure 2.**
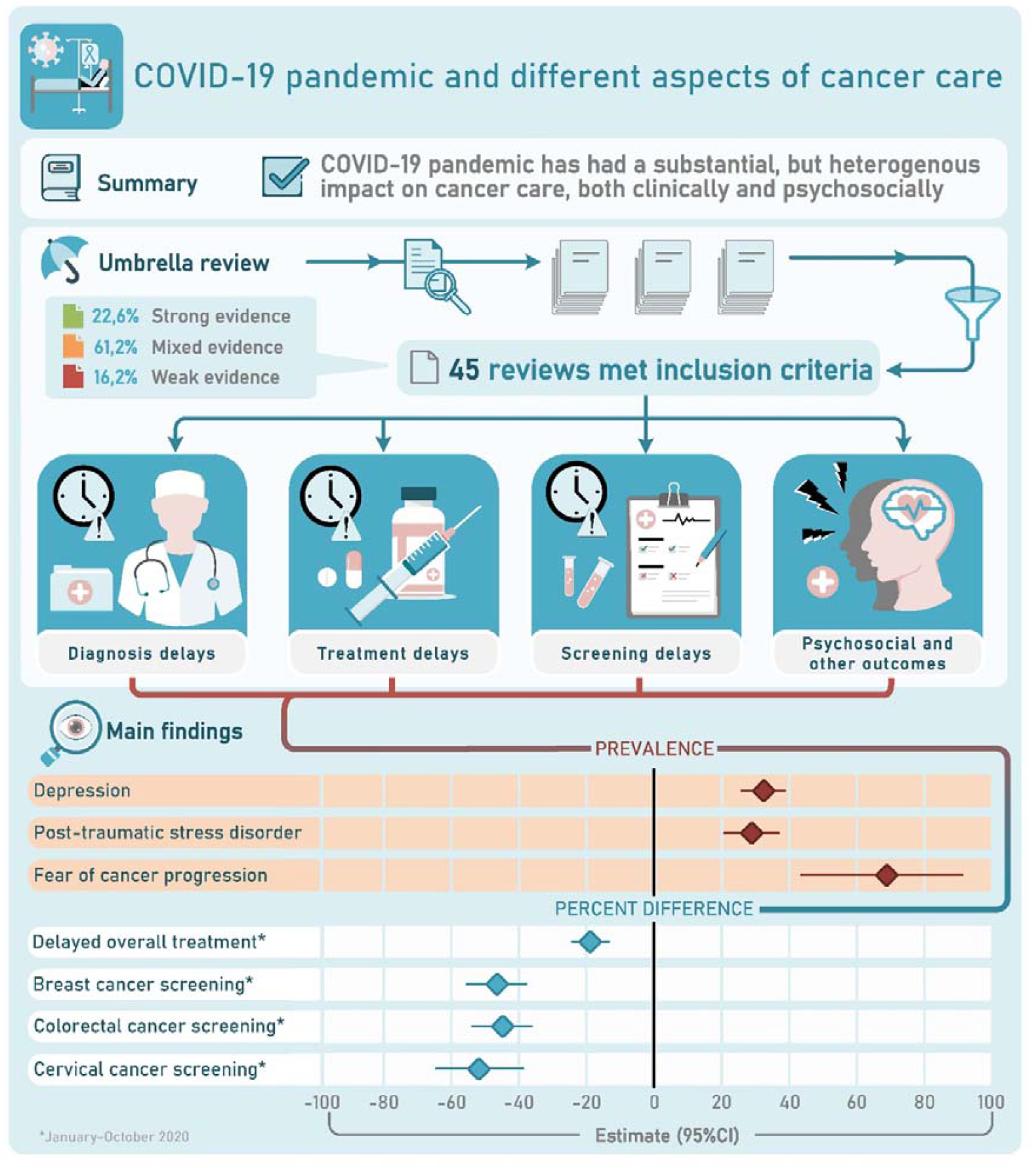
Visual summary.

#### Modification of treatment

There were 11 reviews assessing modification of treatment^5 9 10 18 19 21 26 31 34 37 40^. Main findings for each individual review are outlined in **Appendix 6 and Table 4**. All reviews were consistent reporting changes in treatment, with downscaling treatments plans in cancer patients being a significant intervention. Di Cosimo S et al. 2022 reported changes in treatment plans in 65% (95%CI, 53%-75%; I^2^, 98%) of centers^34^. Guidelines recommended use of non-surgical treatment over surgical treatments, as it was seen in head and neck cancer management. However, reviews suggested patients being assessed in a case-by-case basis and that individual factors should be considered for individualized treatment (**Appendix 6**). Garg PK et al. 2020 found that available guidelines were based on low level of evidence and had significant discordance for the role and timing of surgery, especially in early tumors^21^.

#### Delayed and/or cancelled treatment

**Appendix 7** and **Table 4** summarize the main findings from the 13 reviewes^2 4 5 13 14 22 28 32 34 36 38 40 44^ that assessed and reported on treatment delays and cancellations of cancer treatment. Seven reviews mentioned that cancellations of treatment were observed, although to what extend this happened was not consistently provided^22 28 32 34 36 40 44^. However, reviews reported that these reductions were more pronounced during a lockdown. In the meta-analysis by Teglia F et al., 2022, it was found an overall reduction of −18.7% (95% CI, −13.3 to −24.1) in the total number of cancer treatments administered during January-October 2020 compared to the previous periods, with surgical treatment having a larger decrease compared to medical treatment (−33.9% versus −12.6%); among cancers, the largest decrease was observed for skin cancer (−34.7% [95% CI, −22.5 to −46.8])^4^. This difference would depend on the period, with the review reporting a U-shape for the period January–October 2020.

Lignou S et al. 2022^38^ reported that between 18^th^ to 31^st^ of January 2021, pediatric and noncancer surgical activities were occurring at less than a third of the rate of the previous year, while Di Cosimo SD et al. 2022^34^ reported cancellation/delays of treatment in 58% (95%CI, 48%-67%; I^2^, 98%) of centers. Majeed A et al., 2022^14^ showed that shortage of treatment and delays and interruptions to cancer therapies in general were more common in low- and middle-income countries.

#### Delayed and/or cancelled screening

The results of 11 reviews^3 33 35-37 39 41 42 46 49 51^ reporting on cancer screening are summarized in **Appendix 8** and **Table 4**. Of these, 5 included a meta-analysis. Overall, reviews showed a decline in screening rates across all cancer types, and that differences by demographic area and time periods were observed; for instance, countries that implemented lockdowns showed a higher decline in screening rates. Within colorectal and gastric cancers, most reviews reported a reduction of at least 50% in number of endoscopies and gastroscopies compared to previous years. In the meta-analysis **by** Teglia F et al^3^., while colorectal screening on average was reduced by 44.9% (95% CI, -53.8% to - 36.1%) during January-October 2020, a U-shape association was observed. Within women-specific cancers, the meta-analyses showed a decrease in breast and cervical cancers screening rates of at least 40-50%.^3^ A meta-analysis focused on cytopathology practice showed that on average there was a sample volume reduction of 45.3% (range, 0.1%-98.0%), although the results would depend on the tissue sampled^49^. Similar findings were reported by Alkatoul et al. 2021^33^.

#### Reduced cancer diagnosis

Main findings of the ten reviews^36^ providing data on reduction in cancer diagnosis are provided in **Appendix 9** and **Table 4**. Reviews were consistent in reporting decreased diagnosis of new cancer cases during the pandemic, although the reduction depended on the geographical area, the period being investigated and type of cancer. For example, there was a 73.4% decrease in cervical cancer diagnoses in Portugal during 2020, and in Italy, while there was up to 62% reduced diagnosis of colorectal cancer in 2020 compared to pre-pandemic years, the reduction was more pronounced in Northern Italy where strict lockdowns were implemented. Indeed, reviews showed that countries that implemented lockdowns measures showed the highest reduction in number of new cancer cases being diagnosed. Breast cancer diagnosis rates dropped by an estimate between 18-29% between 2019 and 2021^42^.

#### Reduced uptake of HPV vaccination

There was only one review to summarize data on HPV vaccination, showing up to 96% reduction in number of vaccine doses administered in March-May 2020 among adolescents and young girls aged 9-26 years; the one-year period reduction reported was much smaller (13%)^36^.

#### Psychological needs/distress

Twelve reviews covered topics related to psychological needs and distress that cancer patients experienced during the pandemic^2 5-7 11 20 22 24 27 29 32 37^; the findings are summarized in **Appendix 10** and **Table 4**. Reviews reported that the pandemic negatively impacted the psychosocial and physical wellbeing of cancer survivors and cancer patients experienced different levels of anxiety, depression, and insomnia. In a meta-analysis, Ayubi E et al. 2021 reported an overall prevalence of depression and anxiety of 37% (95%CI, 27-47, I^2^, 99.05) and 38% (95%CI, 31-46%, I^2^, 99.08) in cancer patients, respectively^20^. Similar findings were reported by Zhang et al. 2022^6^. Compared to controls, cancer patients had higher anxiety level [standard mean difference (SMD 0.25 (95% CI, 0.08, 0.42)]^20^.

#### Telemedicine

Telehealth was investigated and reported in 10 of the included reviews^2 10 12 19 23 25 30 32 34 38^; a summary of main findings is provided in **Appendix 11**. Salehi F et al. 2022^30^ reported that telemedicine use in breast cancer patients was the most common investigated in studies exploring cancer-specific use of telemedicine. Telemedicine was used for various reasons, with provision of virtual visit services and consultation being the most common^30^. One study explored various symptom tracking apps for cancer patients, available in the mobile health market, and found that only a limited number of apps exist for cancer-specific symptom tracking (27%)^23^. In addition, of the 41 apps found, only one was tested in a clinical trial for usability among patients with cancer^23^. While little research exists on how patients perceived telemedicine during the COVID-19 pandemic, early data showed that majority of patients found telemedicine service helpful and that obtaining a telemedicine service helped solve their health problem. Nevertheless, there were concerns that use of telehealth for people with cancer suggests a greater proportion of missed diagnoses^38^, and that telemedicine cannot be a substitute for face-to-face appointments^25^.

#### Financial distress and Social isolation

Four reviews reported the economic impact of COVID-19 and social isolation of cancer patients during the pandemic (**Appendix 12**)^2 7 11 22^. While there is little research on this topic, overall, the reviews suggested financial distress with direct and indirect costs burden and social isolation being a common issue for cancer patients. Reviews also were consistent in reporting social isolation and loneliness among cancer patients. Several factors contributed to social isolation, including fear of infection, social distancing measures, not having visitors and lack of social interaction during treatment.

#### Tobacco use and cessation

There was only one systematic review and meta-analysis to explore tobacco use and cessation during the pandemic^45^. Based on data from 31 studies, Sarich P et al. 2022 found that, compared to pre-pandemic period, the proportion of people smoking during the pandemic was lower (pooled prevalence ratio of 0·87 (95%CI:0·79-0·97). In addition, there was similar proportions among smokers before pandemic who smoked more or smoked less during the pandemic, and on average 4% (95%CI: 1-9%) reported stopping smoking. 2% reported starting smoking during the pandemic. High heterogeneity was observed across the meta-analyses results.

#### Other aspects of cancer care

Sixteen reviews^8-10 13 14 18 19 26 28 29 34 38 43 47 48^ reported on mitigations strategies and cancer service restructuring, impact of measures on cancer prognosis, and on quality of recommendations provided during COVID-19 for cancer care; findings are summarized in **Appendix 13**. In the meta-analysis by Di Cosimo S et al., routine use of PPE by patient and healthcare personnel was reported by 81% and 80% of centers, respectively; systematic SARS-CoV-2 screening by nasopharyngeal swabs was reported by only 41% of centers^34^. Four reviews also reported on potential impact of mitigation strategies on cancer outcomes/prognosis^33 38 43 47^. It was estimated that 59,204–63,229 years of life lost might be attributable to delays in cancer diagnosis alone because of the first COVID-19 lockdown in the UK, albeit the findings were based on single study. Delayed cancer screening was estimated to cause globally the following additional numbers of cancer deaths secondary to breast, esophageal, lung, and colorectal cancer, respectively: 54,112–65,756, 31,556–32,644, 86,214–95,195, and 143,081–155,238^33^. Tang et al. 2022^47^ found no deterioration in the surgical outcomes of colorectal cancer surgery or reduction in the quality of cancer removal. Similar findings were also reported by Pararas N et al. 2022^43^, despite the number of patients presenting with metastases during the pandemic was significantly increased. Thomson JD et al. 2020^48^, by exploring recommendations for hypofractionated radiation therapy, found that in general the recommendations during the pandemic were based on lower quality of evidence than the highest quality routinely used dose fractionation schedules.

## DISCUSSION

The current umbrella review appraises systematically the evidence on the extent to which several aspects of cancer care were disrupted during the COVID-19 pandemic. The summary message provided by 45 systematic reviews is that there have been modifications, delays and cancellation of treatment, delays and cancellation in cancer screening and diagnosis, and cancer patients may have experienced additional psychological, social, and financial distress. Nevertheless, appraisal of the impact of COVID-19 on cancer care is mainly based on limited and low-quality evidence, and that data mainly derive from high-income countries, with little understanding of consequences of COVID-19 on cancer care in low- and-middle income countries. In addition, limited evidence exists on whether disruptions in cancer care during the pandemic had adverse impact in prognosis of cancer patients and mortality.

Several guidelines were provided for cancer care during the pandemic, including recommendations on mitigation strategies to prevent SARS-CoV-2 infection and cancer treatment modalities. Nevertheless, most recommendations were based on expert opinions, and little quantitative evidence was provided to support them. This aspect was highlighted also in the systematic review by Thomson JD et al. 2020^48^. The authors explored recommendations for hypofranctionated radiation therapy before and during pandemic and found that during the pandemic there was a significant shift from established higher-quality evidence to lower-quality evidence and expert opinions for the recommended hypofractionated radiation schedules. Similar findings were reported also by Garg PK et al. 2020^21^, suggesting not only guidelines were based on low level of evidence, but also there was significant discordance for the role and timing of surgery, especially in early tumors.

Specific recommendations established from the guidelines such as prioritization of high-grade malignancy, as well as other aspects such as lockdowns, social restrictions, restructure of cancer care with prioritization of high-risk malignancies and use of telemedicine, fear of infection, financial distress and shortage in medications could explain the delays and cancellation in cancer treatment, screening and diagnosis reported in several studies. For example, Lignou S et al. 202^38^ raised concerns that use of telehealth for people with cancer suggests a greater proportion of missed diagnoses. Most of examined systematic reviews reported a substantial reduction in treatment, screening, and diagnosis of several cancers during the pandemic, which was more pronounced for countries that implemented a lockdown. In addition, differences were observed by geographical area, suggesting that the impact on cancer treatment, screening and diagnosis could depend on mitigation strategies countries implemented as well as on country-specific health care organization and resources. For example, shortage of treatment and delays and interruptions to cancer therapies in general were more pronounced in low- and middle-income countries^14^. The findings on disruption of cancer treatment, screening and diagnosis are in line with findings reported for other chronic diseases, such as cardiovascular disease^52^, suggesting the adverse impact might not be cancer specific. Future research should explore and compare how different chronic diseases were impacted.

Evidence is limited on evaluating how disruption of cancer care during COVID-19 affected prognosis of cancer patients. Limited evidence showed that the number of patients presenting with metastases during the pandemic was significantly increased, and emergency presentations and palliative surgeries were more frequent during the pandemic^43^. No deterioration in the surgical outcomes of colorectal cancer surgery including mortality or reduction in the quality of cancer removal was observed^43 47^. A study^53^ in UK estimated that 59,204–63,229 years of life lost might be attributable to delays in cancer diagnosis alone because of the first COVID-19 lockdown, but estimates were based on modelling. Several studies^54 55^ have shown a decline in elective cancer such as colorectal cancer, despite findings showing that gastrointestinal cancer surgery during pandemic is safe with appropriate isolation measures and no delays should be implemented for both early and advanced cancer^56^. A recent meta-analysis^57^ showed that delaying colorectal cancer longer than 4 weeks could be associated with poorer outcomes.

Several studies and systematic reviews thereof have investigated the impact of the pandemic on psychological wellbeing, financial distress, and social isolation of cancer patients, as well as the role of telemedicine in cancer care. While studies suggested depression, anxiety, post traumatic disorder, insomnia and fear of cancer progression being highly reported by cancer patients with estimates reaching beyond 50%, high heterogeneity was observed, and in general systemic analysis comparing the findings with pre-pandemic period rates was lacking. The pandemic was reported to have financial burden on cancer patients with direct and indirect costs. Social isolation was commonly reported and mainly driven by fear of infection, social distancing measures and lack of social interaction during treatment. Nevertheless, there was limited effort to quantify social isolation and economic impact on cancer care. Telemedicine and remote consultations were sharply increased in use for different aspects of cancer care, including treatment, screening, and rehabilitation. However, evidence is limited in evaluating and quantifying the positive and negative impact, as well as cost-effectiveness of telemedicine. While limited evidence suggested telemedicine reduced costs of cancer care for both patients and health care provider, there were concerns especially from patients that telemedicine could not have similar benefits to on-site consultations.

Our study has certain limitations. We searched only one bibliographic database, and therefore we cannot rule out missing some other relevant systematic reviews. Nevertheless, we screened references of included studies to find other relevant studies we may have missed. Most systematic reviews included in this umbrella review were based on intermediate and high risk of bias studies, and the findings were mainly based on case-series, cross-sectional and retrospective observational study designs which are prone to residual confounding and poor in determining temporal associations. Prevalence and incidence estimates are also subject to selection biases. In some instances, data were derived from one study or from studies with small sample sizes and limited number of events, leading to large uncertainty. Many studies did not include any pre-pandemic controls. Furthermore, some of the evidence overlapped among the systematic reviews that were included in this umbrella review, but this allows comparing notes on results and conclusions for the overlapping efforts. Some systematic reviews were published early (in 2020), and thus they had even more limited evidence and the impact of the disruptions may have differed across different pandemic waves. Most findings were derived from high-income and/or western countries, limiting the generalizability of the findings to low- and middle-income countries. Lastly, concreate conclusions on intermediate, and long-term impact remain unclear. Finally, the suboptimal methodological rigor of many included reviews is notable.

In summary, evidence shows a diverse and substantial impact of the COVID-19 pandemic on cancer care, but large uncertainty and gaps exist in the literature on this topic. Future high-quality studies and properly performed, rigorous systematic reviews with careful meta-analyses will continue to have value in this field.

## Data Availability

All data produced in the present work are contained in the manuscript and in the supplemental material.

## Availability of data and materials

All relevant data are included in the manuscript and supplemental material.

## Authors’ Contributions

All authors listed have made a substantial, direct, and intellectual contribution to the work, as well as approved it for publication. T.M., and J.P.A.I., are the guarantors of the work.

