## Appendix 1-3, and 4-13 for "Changes in cancer prevention and management and patient needs during the COVID-19 pandemic: An umbrella review of systematic reviews"

**Appendix 1**. Preferred Reporting Items for Overviews of Reviews- PRIOR Checklist.

**Appendix 2**. Search Strategy

**Appendix 3**. Characteristics of the included reviews

**Appendix** **4**. Bibliographic databases used from each review (see excel file)

**Appendix** **5**. Methodological rigor of included reviews

**Appendix 6**: Summary of results on treatment modification during Covid-19 pandemic

**Appendix** **7**: Summary of results on delays and/or cancellation of cancer treatment during Covid-19 pandemic

**Appendix 8**: Summary of results on delays and/or cancellation of cancer screening during Covid-19 pandemic

**Appendix 9**: Summary of results on delays and/or reduced cancer diagnoses during Covid-19 pandemic

**Appendix 10**: Summary of results on psychological distress of cancer patients during Covid-19 pandemic

**Appendix 11**: Summary of results on telemedicine in cancer care during Covid-19 pandemic

**Appendix 12**: Summary of results on financial distress and social isolation of cancer patients during Covid-19 pandemic

**Appendix 13**: Summary of results on other aspects of cancer care during Covid-19 pandemic

**Appendix 1**. Preferred Reporting Items for Overviews of Reviews- PRIOR Checklist.

| Section topic | Item No | Item | **Location where item is reported** |
| --- | --- | --- | --- |
| Title | 1 | Identify the report as an overview of reviews. | 1 |
| **Abstract** | | |  |
| Abstract | 2 | Provide a comprehensive and accurate summary of the purpose, methods, and results of the overview of reviews. | 2 |
| **Introduction** | | |  |
| Rationale | 3 | Describe the rationale for conducting the overview of reviews in the context of existing knowledge. | 3 |
| Objectives | 4 | Provide an explicit statement of the objective(s) or question(s) addressed by the overview of reviews. | 3-4 |
| **Methods** | | |  |
| Eligibility criteria | 5a | Specify the inclusion and exclusion criteria for the overview of reviews. If supplemental primary studies were included, this should be stated, with a rationale. | 3-4 |
|  | 5b | Specify the definition of “systematic review” as used in the inclusion criteria for the overview of reviews. | 4 |
| Information sources | 6 | Specify all databases, registers, websites, organisations, reference lists, and other sources searched or consulted to identify systematic reviews and supplemental primary studies (if included). Specify the date when each source was last searched or consulted. | 3 |
| Search strategy | 7 | Present the full search strategies for all databases, registers and websites, such that they could be reproduced. Describe any search filters and limits applied. | Supplemental Material |
| Selection process | 8a | Describe the methods used to decide whether a systematic review or supplemental primary study (if included) met the inclusion criteria of the overview of reviews. | 4 |
|  | 8b | Describe how overlap in the populations, interventions, comparators, and/or outcomes of systematic reviews was identified and managed during study selection. | 4-5 |
| Data collection process | 9a | Describe the methods used to collect data from reports. | 4-5 |
|  | 9b | If applicable, describe the methods used to identify and manage primary study overlap at the level of the comparison and outcome during data collection. For each outcome, specify the method used to illustrate and/or quantify the degree of primary study overlap across systematic reviews. | 4-5 |
|  | 9c | If applicable, specify the methods used to manage discrepant data across systematic reviews during data collection. | 4-5 |
| Data items | 10 | List and define all variables and outcomes for which data were sought. Describe any assumptions made and/or measures taken to identify and clarify missing or unclear information. | 4-5 |
| Risk of bias assessment | 11a | Describe the methods used to assess risk of bias or methodological quality of the included systematic reviews. | 4-5 |
|  | 11b | Describe the methods used to collect data on (from the systematic reviews) and/or assess the risk of bias of the primary studies included in the systematic reviews. Provide a justification for instances where flawed, incomplete, or missing assessments are identified but not reassessed. | 4-5 |
|  | 11c | Describe the methods used to assess the risk of bias of supplemental primary studies (if included). | 4-5 |
| Synthesis methods | 12a | Describe the methods used to summarise or synthesise results and provide a rationale for the choice(s). | 5 |
|  | 12b | Describe any methods used to explore possible causes of heterogeneity among results. | 5 |
|  | 12c | Describe any sensitivity analyses conducted to assess the robustness of the synthesised results. | 5 |
| Reporting bias assessment | 13 | Describe the methods used to collect data on (from the systematic reviews) and/or assess the risk of bias due to missing results in a summary or synthesis (arising from reporting biases at the levels of the systematic reviews, primary studies, and supplemental primary studies, if included). | 5 |
| Certainty assessment | 14 | Describe the methods used to collect data on (from the systematic reviews) and/or assess certainty (or confidence) in the body of evidence for an outcome. | 5 |
| **Results** | | |  |
| Systematic review and supplemental primary study selection | 15a | Describe the results of the search and selection process, including the number of records screened, assessed for eligibility, and included in the overview of reviews, ideally with a flow diagram. | 5 |
|  | 15b | Provide a list of studies that might appear to meet the inclusion criteria, but were excluded, with the main reason for exclusion. | 5, Figure 1 |
| Characteristics of systematic reviews and supplemental primary studies | 16 | Cite each included systematic review and supplemental primary study (if included) and present its characteristics. | Table 1 |
| Primary study overlap | 17 | Describe the extent of primary study overlap across the included systematic reviews. | Not provided |
| Risk of bias in systematic reviews, primary studies, and supplemental primary studies | 18a | Present assessments of risk of bias or methodological quality for each included systematic review. | 6 |
|  | 18b | Present assessments (collected from systematic reviews or assessed anew) of the risk of bias of the primary studies included in the systematic reviews. | 5-6, Table 2, Supplemental Material |
|  | 18c | Present assessments of the risk of bias of supplemental primary studies (if included). | 5-6, Supplemental Material |
| Summary or synthesis of results | 19a | For all outcomes, summarise the evidence from the systematic reviews and supplemental primary studies (if included). If meta-analyses were done, present for each the summary estimate and its precision and measures of statistical heterogeneity. If comparing groups, describe the direction of the effect. | 6-9, Table 2-4, Supplemental Material |
|  | 19b | If meta-analyses were done, present results of all investigations of possible causes of heterogeneity. | NA |
|  | 19c | If meta-analyses were done, present results of all sensitivity analyses conducted to assess the robustness of synthesised results. | NA |
| Reporting biases | 20 | Present assessments (collected from systematic reviews and/or assessed anew) of the risk of bias due to missing primary studies, analyses, or results in a summary or synthesis (arising from reporting biases at the levels of the systematic reviews, primary studies, and supplemental primary studies, if included) for each summary or synthesis assessed. | 5 |
| Certainty of evidence | 21 | Present assessments (collected or assessed anew) of certainty (or confidence) in the body of evidence for each outcome. | NA |
| **Discussion** | | |  |
| Discussion | 22a | Summarise the main findings, including any discrepancies in findings across the included systematic reviews and supplemental primary studies (if included). | 10 |
|  | 22b | Provide a general interpretation of the results in the context of other evidence. | 10-11 |
|  | 22c | Discuss any limitations of the evidence from systematic reviews, their primary studies, and supplemental primary studies (if included) included in the overview of reviews. Discuss any limitations of the overview of reviews methods used. | 11-12 |
|  | 22d | Discuss implications for practice, policy, and future research (both systematic reviews and primary research). Consider the relevance of the findings to the end users of the overview of reviews, eg, healthcare providers, policymakers, patients, among others. | 10-12 |
| **Other information** | | |  |
| Registration and protocol | 23a | Provide registration information for the overview of reviews, including register name and registration number, or state that the overview of reviews was not registered. | 1 |
|  | 23b | Indicate where the overview of reviews protocol can be accessed, or state that a protocol was not prepared. | 1 |
|  | 23c | Describe and explain any amendments to information provided at registration or in the protocol. Indicate the stage of the overview of reviews at which amendments were made. | NA |
| Support | 24 | Describe sources of financial or non-financial support for the overview of reviews, and the role of the funders or sponsors in the overview of reviews. | Not provided |
| Competing interests | 25 | Declare any competing interests of the overview of reviews' authors. | 12 |
| Author information | 26a | Provide contact information for the corresponding author. | 1 |
|  | 26b | Describe the contributions of individual authors and identify the guarantor of the overview of reviews. | 12 |
| Availability of data and other materials | 27 | Report which of the following are available, where they can be found, and under which conditions they may be accessed: template data collection forms; data collected from included systematic reviews and supplemental primary studies; analytic code; any other materials used in the overview of reviews. |  |

**Appendix 2**. Search Strategy

***PubMed***

(((cancer) AND (Covid-19 OR SARS-CoV-2 OR COVID19 OR severe acute respiratory syndrome coronavirus 2)) AND (prevention OR screening OR diagnosis OR care OR treatment OR therapy OR management OR financial OR needs OR telehealth OR virtual OR psychological OR social)) AND (systematic review OR meta-analysis)

**Appendix 3**. Characteristics of the included reviews

| **Author, year of publication** | **Systematic reviews/Meta-analysis** | **Searched databases** | **Type of studies considered** | **Type of studies included** | **Number of included studies** | **Covid-19 period covered** | **Risk of bias checklist** | **Methodological rigor / Risk of bias conclusion** | **Pre-pandemic controls** | **Location** | **Cancer type** | **Outcome** | **Analysis** | **Heterogeneity** |
| --- | --- | --- | --- | --- | --- | --- | --- | --- | --- | --- | --- | --- | --- | --- |
| Adham M et al. 2022 | SR | 3 (PubMed, Cochrane, and Clinical Key | Any design except for comments, letters, and case reports | Literature revies and guidelines | 5 | Until July 15^th^ 2020 | Critical appraisal tool of qualitative studies from Centre of Evidence-based Medicine (CEBM), University of Oxford | NP | NS | Globally | Head and neck cancer | Modification of treatment | Qualitative | No |
| Alom S et al., 2021 | SR | 2 (Global Health Medline and EMBASE) | Any design | Any design | 72 | September 2020 | NIH quality assessment tool | NP | NS | Globally, mainly from high-income/upper-middle income countries | Any cancer type, and per different types | - Management and treatment of patient - Reduced diagnosis - Psychological distress - Use of telehealth | Qualitative | No |
| Ayubi E et al. 2021 | SR and MA | 3 (PubMed, Scopus, and Web of Science) | Observational study design, excluding review, editorial, commentary and case-reports | Cross-sectional, survey, case-control study, longitudinal, before-after study | 34 | January 3^rd^, 2021 | NA | NP | No | Globally, 1/3 from China | Any cancer type | Psychological distress   - Depression - Anxiety | - Meta-analysis (DerSimonian-Laird method) of - proportions (metaprop), - standardized mean difference - Subgroup analyses | I2 statistic, high heterogeneity |
| Dhada S et al. 2021 | SR | 5 (CINAHL, EMBASE, MEDLINE, Google Scholar and Wellcome Open Research Authorea) | Any study design | Cross-sectional survey, qualitative studies | 19 | December 2020 | CASP tool for quality studies and NIH quality of Observational, Cohort and Cross-sectional Studies Assessment Tool for survey studies | Mixed/intermediate | No | 10 countries, Italy, S, UK, Netherlands | Any cancer type | - Delayed cancer screening and treatment - Financial, social and psychological distress | Qualitative | No |
| Donkor et al. 2021 | SR | 3 (Ovid Embase, Ovid Medline and CINAHL) | Any design | Case report, expert consensus, cross-sectional survey and critical review | 11 | August 3^rd^, 2020 | Joanna Briggs Institute Critical Appraisal Checklist for Qualitative research | Weak | No | 4 countries, LMIC including China, Iran, Brazil and Zambia | Any cancer type | - Management and treatment of patient - Use of telehealth | Qualitative | No |
| Garg PK et al. 2020 | SR | 3, Medeline, Embase and Scopus | Any design | Guidelines/recommendations/review, research article/survey, case reports/series, editorials, and short communication/commentary/expert opinions | 212 | May 2^nd,^ 2020 | NA | Low level of evidence | NS | Globally, majority of data originated from  countries like the United States, China, and Italy | Any cancer type; data on 12 types of cancer | - Management and treatment of patient | Qualitative | No test, there is significant  heterogeneity |
| Gascon L et al. 2020 | SR | 4, PubMed, Google Scholar, Ovid Medline and Scopus | Guidelines and recommendation documents | Guidelines | 23 | May 2020 | The Agree II (Appraisal of Guidelines for Research and Evaluation II) | Twenty out of the 23 guidelines showed an overall appreciation score of 6 and above. The mean scores (range; SD) for the domains were the  following: scope and purpose 90.8% (16.7–100%; SD 18.2);  stakeholder involvement 70.2% (16.7–100%; SD 19.9); rigor  of development 52.1% (21.9–74.0%; SD 12.6); clarity  of presentation 92.0% (63.9–100%; SD 10.8); applicability  77.4% (56.3–100%; SD 12.5) and editorial independence  26.9% (0–100%; SD 38.3). | No | Globally, North America, South America, Europe, Asia and Oceania | Head and Neck cancer | - Management and treatment of patient | Qualitative | No test, not specified |
| Hojaij FC et al. 2020 | SR | 3, PubMed, Scientific Electronic Library and Scopus | Any design | Editorial, original article, review, case report, opinion/commentary, letters | 35 | Year of 2020 | NA | NP | No | Globally (not clearly specified) | Head and Neck, and otorhinolaryngology | - Use of telemedicine - Management and treatment of patient | Qualitative | No test, not specified |
| Jammu AS et al. 2021 | SR | 3, PubMed, Scopus and Google Scholar | Any design | Expert opinions, literature reviews, prospective cohort studies, retrospective cohort studies, descriptive study and pooled meta-analysis | 19 | August 27^th^, 2020 | NA | NP | NS | Globally, USA, Canada, Hong Kong, Australia, China, Srin Lanka and multi collaboration | Any cancer | - Delays in treatment - Physiological wellbeing of cancer survivors - Economic consequences among cancer survivors | Descriptive | No test, no specification |
| Kirby A et al. 2022 | SR | 5, CINAHL, Medline, PsycINFO, PsycArticles and Embase | Patient perspective, observationnel, cross-sectional, prospective and retrospective studies | Patient perspective, observationnel, cross-sectional, prospective and retrospective studies | 56 | March 31^st,^ 2021 | Joanna Briggs Institute Critical Appraisal Tool and Consensus on Health Economic Criteria (CHEC). Risk of bias in a study was considered  high if the “yes” score was ≤ 4; mode risk  if the score was ≥ 7 on the JBI tools. | Based on JBI tool: 3 at high risk of bias; 23 at moderate risk of bias; 24 at low risk of bias |  | Globally, mainly from USA, Italy, India and China. | Any cancer type, providing data for 7 cancer-specific sites | - Physiological consequences - Social consequences - Economic consequences | Qualitative | No test, no specification |
| Legge H et al. 2022 | SR | 3, CINAHL, Medline and APA PsycINFO | Any study design | Qualitative, quantitative, mixed methods | 18 | February 25^th^, 2022 | Mixed Methods Appraisal Tool (MMAT) | Overall good quality: only three studies were at high or medium risk of bias |  | Globally, mainly from USA and Europe | Any cancer type, 4 cancer-specific information | - Supportive Care Needs (physical, psychological, family related, social and other needs) | Qualitative | No test, insights  across heterogenous study populations in terms experiences  of unmet supportive care needs experienced during  COVID-19 |
| Lu DJ et al. 2021 | SR | 2, iOS App store and Android Google Play | Mobile Health Applications | Mobile Health Applications | 41 apps | May 2020 | Mobile apps rating scale  (MARS) | The app quality mean scores assessed using the mobile apps rating scale ranged from 2.43 to 4.23  (out of 5.00). | No | NA | Any cancer site, and cancer specific  Most apps (30/41, 73%) that met inclusion criteria were  general health/pain symptom trackers, and 11 of 41 (27%) were cancer-specific apps. Of the cancer-specific apps, 5  of 11 (46%) were nonspecific, whereas the remaining 6  included 1 each (1/11, 9%) for blood, lymphoma, head  and neck, breast, pancreatic, and ovarian cancers,  respectively. | - Telehealth - Evaluation of Mobile Health Applications to   Track Patient-Reported Outcomes for Oncology Patients: | Qualitative | NA |
| Momenimovahed Z et al. 2021 | SR and meta-synthesis | 3, PubMed, Web of Science and Scopus | Original study | Cross-sectional, cohort study, qualitative, RCT | 55 | End of June 2021 | NA | NP | NS | Globally, mainly US and Europe | Any cancer site, and cancer-specific information | - Psychological distress | Qualitative | No test, no specification |
| Mostafaei A et al. 2022 | SR and meta-synthesis | 9, MEDLINE (via  Ovid), Embase, Pubmed, Cinahl (via Ebsco), PsycInfo  (via Ebsco), Scopus, ISI Web of Knowledge, Cochrane  Library, and Google Scholar | Qualitative, including phenomenology, ethnography, case studies and grounded theory | Open-ended questions within a cross-sectional quantitative study, mixed-methods design, grounded theory, scenario-based approach | 22 | June 2021 | JBI-Qualitative Appraisal Instrument, a score of seven and above was qualified as high quality | The score varied from 4 to 10, with 16 studies evaluate to be at low risk of bias | No | Globally, mainly in US, Canadian and European context | Any cancer type, and 5 cancer-specific | - Telehealth | Meta-synthesis. Qualitative. Primary author`s interpretations were transferred to MAXQDA software version 10 and further analyzed using open coding to develop categories. Level of credibility was allocated based on risk of bias assessment, and ConQual approach was used to grade the findings. | No test, no specification |
| Moujaess E et al. 2020 | SR | 1, PubMed | Any study design | Mainly short editorials, letters, correspondence or comments, and cohort studies (n=10- retrospective, prospective or cross-sectional analysis) as well as 9 case reports and one case series. | 88 | April 5^th^ 2020 | Not applied | NP | No | Globally, most from China and Italy (52 of 88 studies), US (13%), France (8%), UK (6%) and other countries. | Any cancer site | - Modification of treatment - Other- service restructure/mitigation | Qualitative | No test, no specification |
| Muls A et al. 2022 | SR | 5, EMBASE, Global Health, HMIC, PsychINFO, CINAHL | Qualitative, quantitative, and mixed method primary research, including prospective obestional cohort, cross-sectional | 43 quantiative with a survey method approach, 6 qualitative and 4 used a mixed method design. Primarily cross-sectional cohorts. RCTs, non-randomized controlled/crossover trials, case control studies, case reports, observational studies. | 51 | October 2021 | Mixed Methods Appraisal Tool (MMAT) | Most of the studies received a  medium score, 16 studies (30%) scored four  or five stars.  Majority of papers acquired a score of 2 (15 studies) or 3 (17 studies) out of a maximum 5 score. Limitations included small samples, non-representative samples, lack of detail relating to cancer types and treatment and methodological flaws |  | Globally, 21 countries | Various cancer types (52%, 27 studies), 48 cancer specific. | - Emotional and psychological impact | Meta-synthesis. The  process involved identifying key concepts from studies and translating  them into one another. The term “translating” refers to the  process of extracting concepts from one study and acknowledging  the same concept in another study even if the concepts are  expressed in different words. | No test, studies used many different outcome measures |
| Murphy A et al. 2022 | SR | 5, CINAHL, Medline, PsycINFO, PsycArticles and Embase | Patient perspective,  Observational,  Cross-sectional,  Prospective,  Longitudinal  Retrospective | Prevalence studies (17), cross-sectional analytical studies (4), qualitative studies (2), cohort studies (13), Cost analysis (1). | 37 | March 31st, 2021 | JBI critical appraisal tools, and CHEC list. Risk of bias in a study was considered high if the “yes” score  was 4; moderate if 5–6; and low risk if the score was 7 on the JBI tools | 1 study at high risk of bias, 19 at moderate risk of bias, 16 at low risk of bias |  | Globally, mainly from USA (35%) | Any cancer site, multiple/all cancer types (67%). | - Telehealth, economic, social, psychological and health impact | Qualitative | No test, no specification |
| Pacheco RF et al. 2021 | SR | 10, CINAHL, Cochrane library, Embase (via Elsevier), Epistemonikos, Health Systems Evidence, LILACS, and Medline (via PubMed), McMaster Daily News Covid-19, Oxford Covid-19 Evidence Service and WHO Global Literature on Coronavirus Disease | **Experimental studies** (randomized, quasi-randomized,  and nonrandomized trials; single experimental cohort,  or controlled before-and-after studies), **Observational longitudinal comparative studies** (cohort  or case-control)  • Observational noncomparative studies (case series or  case studies reporting the experience of a specific  cancer service; Cross**-**sectional studies (prevalence, survey, or analytical  cross-sectional); Uncontrolled before-and-after studies (including  interrupted time series studies with two or more  measures before and after the event of interest) | Two case series, three cross-sectional  studies, and four analytical cross-sectional  studies | 9 | NP | Cochrane Risk of Bias Table, ROBINS-I and JBI tool | The methodological quality was considered low (1 study) to moderate (1 study) for case series and low for all cross-sectional and analytical cross-sectional studies. | NS | USA, Italy, China, Spain, UK and Iran | Any cancer site, including breast cancer, head and neck cancer and lung cancer. | - Delays in treatment and diagnosis- administrative and economic outcomes - Management and treatment of patient: mitigation strategies | Qualitative, but quantitative analysis was planned if sufficient and homogenous data were provided | Considering the scarcity and underreporting of data and the  clinical and methodological heterogeneity among included  studies, meta-analyses were not appropriated |
| Rohilla KK et al. 2021 | SR | 5, PubMed, Medline, Embase, Clinical Kew and Google Scholar | Any design | Editorial (1 study), review (2 studies), observational (2 studies), guideline for palliative care | 6 | February 3^rd^, 2021 | Studies were assessed for validity and authenticity by five  experts from different oncology departments | NP | NS | India | Any cancer site | - Quality of life | Qualitative | No test, no specification |
| Salehi F et ak. 2022 | SR | 4, PubMed, Science Direct, Web of Science and Embase | Any study design except for letters to editor, short communications | Case report (1 study), cross-sectional (12 studies), multi method (1 study) and cohort study (2 studies) | 16 | April 2021 | NA | NP | NS | Globally, half from US (8 studies), Canada, Brazil, China, Italy, Turkey | Any cancer site | - Telehealth | Qualitative | No test, no specification |
| Sun P et al. 2021 | SR | 3, PubMed, Cochrane Library and Embase | Any study design | NP | 6 | February 2021 | NA | NP | No | Italy (2 studies), America (2 studies) England (2 studies) | Breast cancer | Management and treatment of breast reconstruction | Qualitative | No test, no specification |
| Zapala J et al. 2022 | SR | 3, Medline (PubMed), Google Scholar and PsycINFO | Any study design | Not provided | 160 | Not provided, and studies not during Covid-19 area were included | NA | NP | NS | NP | Any cancer site | - Psychological, ethical and spiritual aspects - Telehealth - Delays in treatment and diagnosis | Qualitative | No test, no specification |
| Zhang L et al. 2022 | SR and MA | 4, PubMed, Embase, PsycINFO and Web of Science | Cross  sectional or cohort studies. | 36 cross-sectional and 4 cohort studies (data extracted only for one time point) | 40 | January 31^st^, 2022 | The JBI tool | 11, 14 and 15 studies had high, unclear, and low risk of bias, respectively. Risk of bias was relatively small, indicating moderate methodological quality | No | Globally, mainly from China, Italy, Netherlands, America, and Canada | Any cancer | - Psychological disorders (anxiety, depression, PTSD, insomnia, distress, and fear of cancer - progression/recurrence during COVID‐19 pandemic) | Qualitative and quantitative (meta-analysis using random effect model).  To explore the source  of heterogeneity, a subgroup analysis was conducted according to  several variables (area, scale, risk of bias, cancer type, gender,  marital status, education level, and employment status). | ≥ 0.05 and I2 ≤ 50% represented no heterogeneity,  and a fixed‐effects model was applied. High heterogeneity in most analyses |
| AlkatouI et al. 2021 | SR | 4, PubMed/Medline, Scopus and Web of Science | Any observational study design, except for case-reports, letters to editors, commentaries and reports | Prospective (2 studies), retrospective (12 studies), simulation study (2 studies) | 16 | December 28^th^, 2020 | Newcastle-Ottawa Quality Assessment Form | 15 studies good quality and one article moderate quality | Yes | Globally, USA, Taiwan, Belgium, Netherlands, Japan, Italy, England, Austria and Canada | Any cancer site, breast cancer, colorectal cancer, gynecological cancer, lung cancer, colon cancer | - Delayed/cancelled screening - Reduced diagnoses | Qualitative | No test, no specification |
| Cosimo SD et al. 2022 | SR and MA | 3, PubMed, Scopus and Web of Science | Surveys | Surveys | 56 | December 11, 2020 | “Risk of bias instrument for cross-sectional  surveys of attitudes and practices” from the CLARITY Group at  McMaster University | From 1 to 5, all studies (except for one) scored 3 or less, suggesting moderate to high risk of bias. | Yes | Globally, 121 countries | Any cancer site, and cancer-specific including brain, head and neck, gynecological,  breast, hepato-bilio-pancreatic, hematological, colorectal, skin,  pediatric, urinary tract, esophagogastric, neuroendocrine, lung  and soft tissues cancers. | - Cancellation/delay of treatments - Modification of treatments, - Cancellation/delay of clinic visits, reduction of activity - Telehealth | Qualitative and Quantitative.  A random-effects meta-analysis was performed using the  DerSimonian-Laird estimator for variance.  A thorough moderator analysis was  performed to account for this heterogeneity with subgroup  analysis and meta-regression | I2, P- value for the Q-test. High heterogeneity was observed across the meta-analyses |
| Fancellu A et al. 2022 | SR | 4, PubMed, Scopus, Web of Science and Reference Citation Analysis | Observational retrospective studies, surveys, national or regional database-studies | Single-unit or nation-wide surveys | 7 | January 31, 2022 | Not Applied | NP | Yes | Italy | Colorectal Cancer | - Delayed/cancelled screening - Reduced diagnosis | Qualitative | No test, no specification |
| Ferrara P et al. 2022 | SR | 4, PubMed/Medline, Scopus and Embase | Trial or observational studies such as case-control, cohort, or cross-sectional studies | All studies had an observational design | 33 | February 8^th^, 2022 | Newcastle-Ottawa Scale (aNOS), reports achieving an aNOS score of 5 or greater were considered high-quality studies | Methodological quality varied across the 34 studies, of which 32 scored five or more stars on the aNOS quality assessment, while two were classified as low quality given the high risk of bias. Major reasons for bias across studies included lack of representativeness of the sampled participants, as well as substandard assessment of the outcomes as they were mostly self-reported through questionnaires. | Yes | Globally, 16 studies were carried out in America, 13 in Europe, three in Asia, and two in Africa | Cervical cancer | - Reduced uptake of HPV vaccination - Delay/cancellation of treatment - Modification of treatment - Delayed/cancelled screening - Reduced diagnoses | Qualitative | No test, there was high significant heterogeneity in methods and outcomes across the retrieved studies |
| Gadsden T et al. 2022 | SR | 3, Ovid Medline, Embase and Global Health. Additional nonstructured searches for grey literature  were conducted in the WHO COVID-19 database and a  pre-print database (e.g., https://www.medrxiv.org/). | Observational studies | Nearly all studies were cross-sectional in nature, though different study design labels were  applied including ‘ambi-directional’ cohort study and prospective  mixed-quantitative methods study | 17 | December 15, 2021 | JBI, ROBINS-I risk of bias  tool, the Cochrane Collaboration to  assess non-randomized studies of interventions | Moderate to high risk of bias. Most studies outlined clear time periods for comparison, the study setting, outcomes for measurement and their data source. Comparatively, sample size was not reported in few studies, and studies commonly did not explain their statistical analysis methods nor provide confidence intervals. No study controlled for confounding. None of the included studies were scored a high methodological quality. | Yes | Majority conducted in India (13 studies), Indonesia (1 study), Sri Lanka (2 studies), Bangladesh (1 study) | Any cancer site and cancer-specific, including cervical cancer (n=1), pediatric (n=1), oral (n=1), blood (n=1), gastrointestinal (n=1) and head and neck (n=1) cancers | - Delay/cancellation of treatment - Modification of treatment - Reduced diagnoses | Qualitative | No test, there was high heterogeneity in setting, population, condition,  and service area. |
| Hesary FB et al. 2022 | SR | 2, Medline and Web of Science | Observational studies, excluding reviews, case reports, letters to editor and clinical trial | Cross-sectional, retrospective, perspectives, prospective | 22 | 2021 | The Newcastle-Ottawa Quality Assessment Form | Low to moderate risk of bias; 17 studies defined as good quality and 5 studies as fair quality | Yes | Globally, Italy, UK, Portugal, Netherlands, China, India, Japan, Turkey, Iran, Singapore | Gastric Cancer | - Delayed/cancelled screening - Reduced diagnoses - Modification of treatment - Psychological needs | Qualitative | No test, no specification |
| Lignou S et al. 2022 | SR in children | 2, PubMed and PsycINFO | Any study design | Literature review, Survey, retrospective, interviews, population based modelling, multi-center surveillance, | 32, only one specifically to cancer, some on multiple chronic diseases | August 2021 | Not applied | NP | Yes | Globally, the cancer data mainly from UK | Pediatric cancer | - Delay/cancellation of treatment - Modification of treatment - Delayed/cancelled screening - Reduced diagnoses - Telehealth - Other; impact on disease severity, increasing mortality and increasing emergency and hospital admissions | Qualitative | No test  , no specification |
| Majeed A et al. 2022 | SR | 4, PubMed, Embase, CINAHL and the African Medical Index | Any study design except for mechanistic and preclinical studies | Case series, letters, opinion papers, cross-sectional, case series, cohort studies, cross-sectional, qualitative. | 132, 60 evaluating cancer care delivery during pandemic | November 3, 2021 | GRADE | Low to moderate within care delivery studies; 23 very low certainty, 12 low, 8 moderate and one high certainty. For some studies was not possible to do the assemsnet | Yes, but NS | Globally, Italy (6 studies), US (19 studies), France (3 studies), Poland (4 studies), Saudi Arabia (2 studies), Spain (5 studies), Greece (2 studies) and other countries. | Pediatric cancer | - Delay/cancellation of treatment - Modification of treatment - Delayed/cancelled screening - Reduced diagnoses | Qualitative | No test, the methodological heterogeneity amongst  reported studies (e.g., due to patient population, types of  tumors studied; clinical outcomes measured) limited the ability  to quantitatively assess data between cancer diagnoses and  between HICs/LMICs. |
| Mayo M et al. 2021 | SR and MA | 5, PubMed, Ovid Medline, Cochrane Covid-19 study register, ClinicalTrials.gov and Embase | Retrospective observational studies of cohorts or cancer registries | Retrospective observational studies | 13 | February 10^th^, 2021 | Newcastle-Ottawa Quality Assessment Tool and GRADE | Moderate to low risk of bias; 6 studies at low risk of bias, 7 at moderate risk of bias.  Based on GRADE, quality of evidence  to be high for diminished colon cancer  screening and moderate for the diminution of  breast and cervical cancer screening during  the beginning of the COVID-19 pandemic | Yes | Globally, Italy, Australia, Taiwan (3 studies), US (3 studies), France and Netherlands | Any cancer site, but identified mainly   - breast   cancer   - colon cancer - cervical cancer   lung cancer | - Delayed/cancelled screening. | Qualitative and Quantitative  The pooled IRRs were calculated  using the inverse variance method, and  random effects models were presented. Leave-one-out analyses  were performed to calculate pooled estimates  to determine if studies with high  influence was impacting the significance of  the results. | X2 and I2 statistics, high heterogeneity observed across meta-analyses |
| Mazidimoradi A et al.2021 | SR | 4, PubMed/Medline, Scopus and Web of Science | Any observational study design | Retrospective cohort, service evaluation, national survey, cross-sectional, web-based and online survey, research letter, retrospective review, descriptive analysis, ambispective analysis | 43 | June, 2021 | Newcastle-Ottawa Quality Assessment Scale | Low to moderate risk of bias; 22 articles defined as good quality and 16 articles as medium quality, 5 as poor quality | Yes | Globally, most eligible studies have  been conducted in European countries 38 articles as well  as India and China. | Colorectal cancer | - Delay/cancellation of treatment - Modification of treatment - Delayed/cancelled screening - Reduced diagnoses | Qualitative | No test, no specification |
| Mazidimoradi A et al. 2022 | SR | 3, PubMed/Medline, Scopus, and Web of Science | Any observational study design | Retrospective, cross-sectional, service evaluation, national survey, prospective observational, commentary | 25 | June 2021 | Newcastle-Ottawa Quality Assessment Scale | Low to medium risk of bias; 22 articles were defined as good quality and 3 articles as medium quality |  | Globally, mainly from Europe (15 studies). | Colorectal cancer | - Delayed/cancelled screening | Qualitative | No test, no specification |
| Ng JS et al. 2022 | SR and MA | 4, Ovid Medline, Ovid Embase, medRxiv and bioRxiv | Retrospective cohort studies | Retrospective cohort study | 31 | October 1^st^, 2020 | Newcastle-Ottawa Scale (NOS). Studies were graded by a single reviewer as high (8-9), moderate (6-7) or low (≤5) quality based on the number of stars awarded in the assessment scale | Low to moderate risk of bias; 7 studies at low risk of bias, 24 at moderate risk of bias. Most studies lost one or two points in the comparability domain for not accounting for the effects of confounders in the studies | Yes | Globally, US (8 studies), Italy (3 studies), UK (3 studies), Austria (2 studies) and Taiwan (2 studies). | Breast cancer | - Delayed/cancelled screening - Reduced diagnoses | Qualitative and quantitative. Rate ratios from eligible studies were log-transformed then pooled in Review Manager 5.4 using standard inverse-variance random effects meta-analysis according to the method proposed by Der Simonian and Laird.16 The standard error (SE) of the log rate ratio was estimated as outlined in the Cochrane Handbook for Systematic Reviews of Interventions | I^2,^ Tau ^2,^ and X^2^ test; high heterogeneity observed across meta-analyses |
| Nikolopoulos M et al. 2022 | SR | 3, PubMed, Embase and Cochrane Central | Prospective and retrospective studies | One systematic review was identified. Nine studies were retrospective  reviews of cases and five studies were questionnaire  surveys | 15 | February 10^th^, 2021 | Newcastle–Ottawa Scale | Moderate to high risk of bias; The score ranges from 3 to 7, out of 9 maximum points. | Yes, but NS | Globally, three were conducted in the USA, two in  China and one in each of the following countries: Turkey,  Italy, Spain, India, and Austria. The online studies were conducted  in USA (two), India, and two of them globally | Gynecological cancer, endometrial, cervical, ovarian, and vulval cancer | - Delay/cancellation of treatment - Modification of treatment - Delayed/cancelled screening - Reduced diagnoses - Psychological distress | Qualitative | No test, the retrospective nature of the published reviews and the  small cohort of patients included in each study unavoidably  result in different numeric outcomes and do not allow to  draw definite conclusions |
| Pararas N et al. 2022 | SR and MA | 5, MedLine, Scopus, Web of Science and China National Knowledge Infrastructure (CKNI) database and clinicaltrials.gov | Any observational study design, excluding non-clinical studies |  | 10 | NP | Newcastle–Ottawa scale (NOS). Each study is awarded a score of 0 to 9 stars, quantifying its  methodological quality, with studies scoring 7 to 9 being of high quality, studies with  scores from 5 to 6 being of mediocre quality, and studies with a score of 4 or less being of  poor quality. | Low to moderate risk of bias; eight studies had an NOS score of 7 to 9  and were deemed of high methodological quality, and two studies had a score of 5 or 6 and  were deemed of mediocre methodological quality. The median score of the obtained NOS  scores were 7.5 | Yes | Globally, five studies being from east Asia (one from Japan, two from China, and two from Korea) and the remaining five from Europe (two from the United Kingdom, one from Italy, one from Ireland, and one from Serbia). | Colorectal cancer | - Other: impact of delayed diagnosis and health care changes in cancer stage upshifting or adverse treatment outcomes | The random effects  model was a priori selected to calculate ORs, WMDs, 95% Confidence Intervals (CI), and  relevant p-values due to expected clinical heterogeneity in terms of geography, regional  covid prevalence, and the extent of elective service disruption by Covid. Leave-one-out analysis | I^2^, Tau ^2^, and X^2^ test; Moderate to high heterogeneity |
| Riera R et al. 2021 | SR | 10, CINAHL (via EBSCOhost), Cochrane Library (via Wiley), EMBASE (via Elsevier), Epistemonikos, Health Systems Evidence, LILACS (via Biblioteca Virtual em Saude) and Medline (via PubMed), McMaster Daily News Covid-19, Oxford Covid-19 Evidence Service and WHO-Global Literature on Coronavirus Disease. Additional non-structured searches were performed. | Observational longitudinal comparative studies (cohort or case-control), observational noncomparative studies (case series and case studies), cross-sectional studies (prevalence, survey or analytical cross-sectional), controlled before and after studies, and uncontrolled before and after studies. | Case series, longitudinal studies, cross-sectional, analytical cross-sectional and survey | 62 | NP | ROBINS-I, JBI (the studies were  categorized as presenting high quality [scored 7 or 8],  moderate quality [scored 6 or 5], or low quality [scored 4 or  lower]), IH Quality Assessment Tool for Case Series  Studies and Center for Evidence-Based Management’s critical appraisal of a survey (for survey assessment, considering the 12 questions to be answered, at the discretion of the review authors, the studies were categorized as presenting high quality [scored 9-12], moderate quality [scored 5-8], or low quality [scored 4 or lower]). | Moderate to high risk of bias; The methodological quality was considered low for case series, low for longitudinal studies, and moderate to low for cross-sectional studies. Among analytical cross-sectional studies, the quality was considered moderate for and low for the remaining 14. For surveys, the methodological quality was considered moderate for and low for the remaining eight. | Yes | Globally, majority from Italy (30.6%), USA (16.1%), China (9.7%), France (6.5%), UK (4.8%), Canada (3.2%) and the rest with 1.6% each | Any cancer site, and cancer specific including breast, head and neck, urological, colorectal, skin, hematological, gynecological, pediatric, lung, hepatopancreatobiliary, stomach and musculoskeletal. | - Delay/cancellation of treatment - Delays/Reduced diagnoses | Qualitative | No test, we highlight the pronounced clinical and methodological  heterogeneity among the included studies. The studies were  conducted in several countries with various public and private  health systems and policies. Twelve different cancer conditions  were considered, including early-stage and/or metastatic  disease, and the data were collected during distinct pandemic  stages. Substantial heterogeneity is also noted in the reported  outcome measures. |
| Sarich P et al. 2022 | SR and MA | 6, Medline, Embse, PsycINFO, BioRxiv and MedRxiv, and SSRN website | Cross-sectional studies, cohort studies,  and uncontrolled “before-and-during” studies | Majority cross-sectional studies, but also before and during studies. | 44 | November 5^th^, 2020 | ROBINS-I and Risk of Bias checklist for prevalence studies by Hoy Damian et al. 2012 | High risk of bias; all cross-sectional studies were at high risk of bias, the before and during studies were mix, from low to high risk of bias. The two major  sources of bias were selection of participants into the  study, mainly due to non-representative participants or  low response rates, and in the measurement of the outcome  with different methods and/or tools/questions  used before and during the pandemic. Another major source of  bias being that study populations were not representative  of the target population | Yes | Globally, across 24 countries | NA | - Tobacco use and cessation | Qualitative and Quantitative; random effects models were used to pool data. Subgroup analysis was not feasible. Meta-regression was used to assess the relationships between the included outcomes and (1) the severity of COVID-19 outbreaks in the study population and period (number of COVID-19 cases or deaths per capita between the start and end dates of the survey and also from the start date of the pandemic to the end date of the survey), (2) the mean daily stringency index of the national response to the COVID-19 pandemic during the survey | I^2^) and X^2^. Heterogeneity was  high in all meta-analyses and so the pooled estimates should be interpreted with caution (I2>91% and p-heterogeneity<  0¢001). |
| Sasidharanpillai S et al. 2022 | SR and MA | 4, Pubmed/Medline, Scopus, Google Scholar | Observational studies | Retrospective studies | 7 | September 2021 | National Institute of Health Checklist (NHLBI, NIH).  The studies with a minimum score of eight or above,  seven, or five or less than five “Yes responses” were  considered good, fair, and poor quality, respectively. For  cross-sectional and case-control studies, question numbers  1, 2, 3, 4, 5, and 11 were applicable. The responses to the  remaining eight questions (6-10,12,13,14) were marked as  not applicable (NA). Each question was categorized as Yes,  No, others-CD (can-not determine), NA (not applicable),  NR (not reported). The studies with six “Yes” responses  were considered good, and those with four /five were taken  as fair. The studies with less than four “Yes responses”  were considered of poor quality. Two reviewers assessed  the quality of the studies. | Low risk of bias; all  these studies were qualified as good. | Yes | Globally, Slovenia, Italy, Canada, Scotland, Belgium and US | Cervical Cancer | - Delayed/cancelled screening | Qualitative and Quantitative; metaprop package was used based on random-effects model | I^2^ and X^2^. The I2 value  ranging between 0% to 24% indicates consistency. The  I2 values of 25%-49% and 50-74% point toward low and  moderate heterogeneity, respectively. In studies with high  heterogeneity, the I2 value varied between 75%-100%.  A considerable amount of heterogeneity across  the studies were anticipated as the included studies were  mostly observational. High heterogeneity was observed in meta-analyses |
| Tang G et al. 2022 | SR and MA | 3, Embase, Web of Science and PubMed | Cohort and case-control studies | Retrospective case-control, retrospective cohort study, prospective case-control study | 14 | January 12, 2022 | Newcastle-Ottawa Scale | Low risk of bias | Yes | Globally, Turkey, China, UK, Italy, Denmark, Austria, Australia | Colorectal cancer | - Other; impact on health outcomes | Qualitative and Quantitative; The mean differences or  odds ratios (ORs) for individual studies were combined using  a random effects meta-analysis when I² was >50 %. Otherwise,  the fixed-effect model was selected. Leave-one out analysis was performed. | I^2^ and X^2^. Low to moderate heterogeneity observed ins most meta-analyses |
| Teglia F et al. 2022 | SR and MA | 3, PubMed, ProQuest and Scopus | Observational studies and cancer registries | Observational studies and data from cancer registries | 39 | December 12^th^, 2021 | Critical  Appraisal Skills Programme for qualitative research,15 for  a maximum score of 10 points. Studies obtaining less than 7  points were considered inadequate and excluded (no article  was excluded because of a low-quality score). | Low to moderate risk of bias; all studies scored 7 or higher | Yes | Globally, America, Asia, Europe, | Breast cancer, colorectal cancer and cervical cancer | - Delayed/cancelled screening | Qualitative and Quantitative. For each type of screening, the weighted average of the percentage variation between screening tests performed was calculated before and after the beginning of the COVID-19 pandemic. The weight was calculated with the natural logarithm of the number of daily events in the prepandemic period (ie, daily_screen_precovid obtained by dividing the number of screening tests in the pre-pandemic period by its duration in days: weight = ln [daily_screen_precovid]). The logarithm was used because of the great variability in the number of tests between studies. Absolute value was used to avoid negative weights. An ordinary least-squares linear model was fitted using Newton-Raphson (maximum likelihood) optimization with percentage change as dependent variables and terms for type of structure , geographic area, and period as independent variables. P values of differences of means are based on the  t test, those of differences of proportions are based on z scores,  and P tests of multivariate analyses are those derived from  the regression generalized linear models for the respective  variables. | No test, considerable heterogeneity  between countries was present in terms of screening  protocols, services’ accessibility, and participation of the target  population, lockdownmeasures,andincidence ofCOVID-19  and its temporal trend. |
| Teglia F et al. 2022 | SR and MA | 3, PubMed, Proquest and Scopus | Observational studies and cancer registries | Observational studies and data from cancer registries | 47 | December 12, 2021 | Critical Appraisal Skills Programme (CASP) | Low to moderate risk of bias; all studies scored 7 or higher | Yes | Globally | Any cancer and cancer-specific | - Delayed/ cancelled treatment | Qualitative and quantitative; weighted average for the number of daily events. Subgroup analyses were performed by geographical area, type of setting and period | No test |
| Thomson JD et al. 2020 | SR | 2, PubMed/Medline and websites of national and international organizations | Published recommendations related to dose fractionation | Recommendations | 54 | June 1, 2020 | The American Society of Radiation Oncology (ASTRO) classification The ASTRO scale defines 4 levels of quality of evidence: high, moderate, low, and expert opinion. To be designated high quality, the fractionation schedule had to be supported by 2 or more well conducted and highly generalizable randomized clinical trials or meta-analyses of such trials | Mixed quality, cancer-specific results. In general, low to moderate quality. | Yes | Not provided | Hypofractionated radiation therapy for any cancer sit, and cancer-specific including breast, central nervous system, cutaneous, nonmelanoma, upper gastrointestinal, lung, lower gastrointestinal, genitourinary, gynecology, head and neck, hematologic, lymphoma, pediatrics, general palliative and sarcoma | - Other; quality of recommendations | Qualitative and Quantitative. Contingency tables with c2 tests were used to  evaluate the distribution of the quality of evidence of the  highest-rated schedules compared with that of the COVID era  schedules. Analysis of variance methods were used to  determine differences between disease groups. Scatter regression plots were used to visualize the  overall changes in quality from the highest-quality schedules  for specific clinical scenarios to the quality of evidence  of the alternative schedules proposed in the pandemic-era  literature. The shifts in the quality of evidence from “pre-COVID”  to the highest-ranked “in-COVID” site-specific recommendations  were compared. The disease sites with less  substantial shifts were compared with those with greater  changes in quality using the adjusted c2 test. Differences  between disease sites were further compared using a  weighted shift based on the “pre-COVID” evidence quality  and the levels of evidentiary shift to the “in-COVID”  ranking, with significance determined by the adjusted c2  test. The weights were assigned according to a progressive  hierarchy of the shifts high to opinion, high to low, high  to moderate, moderate to opinion, moderate to low, and low  to opinion receiving a numerical value from 6 to 1,  respectively. Top-weighted shifts were compared with lowweighted  shifts around the median | No tests, there was heterogeneity in levels of evidence provided |
| Vigliar E et al., 2020 | Individual-participants meta-analysis | CytoESP Working Group (cytopathologists from the European Society of Pathology) (https://www.esp-patho logy.org/worki nggroups/esp-worki ng-group s/cytop athol ogy.html) and to cytopathologists who have taken part in 1 of the 9 Annual National Molecular Cytopathology meetings in Naples, Italy (https://www.molec ularc ytopa tholo gy.com/), accounting for a total of 65 invited participants. | Survey | Survey | 41 respondents | April 30, 2020, | Not applicable | Not applicable | Yes | Globally, 23 countries | Any cancer, Cytopathology Practice | - Delayed/cancelled screening - Delays/Reduced diagnoses | Qualitative and Quantitative; The random effects model of DerSimonian  and Laird was a priori selected due to the anticipated heterogeneity  among institutions. Global differences between the 2 periods with respect  to the percentage of samples for each single anatomic  site were assessed using the Fisher exact test and  the corresponding P values were adjusted for multiplicity  using the Benjamini-Hochberg correction procedure | I^2^, tau^2^ and X^2^. not due to sampling error). Standard thresholds were considered for the determination of I²: ≤25%  for low heterogeneity, 26% to 50% for moderate heterogeneity,  and >50% for high heterogeneity. High heterogeneity in most analyses. |

Appendix 5. Methodological rigor of included reviews

| **Author, year of publication** | **Number of included studies** | **Risk of bias checklist** | **Methodological rigor / Risk of bias conclusion** | **Pre-pandemic controls** | **Location** |
| --- | --- | --- | --- | --- | --- |
| Adham M et al. 2022 | 5 | Critical appraisal tool of qualitative studies from Centre of Evidence-based Medicine (CEBM), University of Oxford | NP | NS | Globally |
| Alom S et al., 2021 | 72 | NIH quality assessment tool | NP | NS | Globally, mainly from high-income/upper-middle income countries |
| Ayubi E et al. 2021 | 34 | NA | NP | No | Globally, 1/3 from China |
| Dhada S et al. 2021 | 19 | CASP tool for quality studies and NIH quality of Observational, Cohort and Cross-sectional Studies Assessment Tool for survey studies | Mixed/intermediate | No | 10 countries, Italy, S, UK, Netherlands |
| Donkor et al. 2021 | 11 | Joanna Briggs Institute Critical Appraisal Checklist for Qualitative research | Weak | No | 4 countries, LMIC including China, Iran, Brazil, and Zambia |
| Garg PK et al. 2020 | 212 | NA | Low level of evidence | NS | Globally, majority of data originated from  countries like the United States, China, and Italy |
| Gascon L et al. 2020 | 23 | The Agree II (Appraisal of Guidelines for Research and Evaluation II) | Twenty out of the 23 guidelines showed an overall appreciation score of 6 and above. The mean scores (range; SD) for the domains were the  following: scope and purpose 90.8% (16.7–100%; SD 18.2);  stakeholder involvement 70.2% (16.7–100%; SD 19.9); rigour  of development 52.1% (21.9–74.0%; SD 12.6); clarity  of presentation 92.0% (63.9–100%; SD 10.8); applicability  77.4% (56.3–100%; SD 12.5) and editorial independence  26.9% (0–100%; SD 38.3). | No | Globally, North America, South America, Europe, Asia and Oceania |
| Hojaij FC et al. 2020 | 35 | NA | NP | No | Globally (not clearly specified) |
| Jammu AS et al. 2021 | 19 | NA | NP | NS | Globally, USA, Canada, Hong Kong, Australia, China, Srin Lanka and multi collaboration |
| Kirby A et al. 2022 | 56 | Joanna Briggs Institute Critical Appraisal Tool and Consensus on Health Economic Criteria (CHEC). Risk of bias in a study was considered  high if the “yes” score was ≤ 4; mode risk  if the score was ≥ 7 on the JBI tools. | Based on JBI tool: 3 at high risk of bias; 23 at moderate risk of bias; 24 at low risk of bias | No | Globally, mainly from USA, Italy, India, and China. |
| Legge H et al. 2022 | 18 | Mixed Methods Appraisal Tool (MMAT) | Overall good quality: only three studies were at high or medium risk of bias | No | Globally, mainly from USA and Europe |
| Lu DJ et al. 2021 | 41 apps | Mobile apps rating scale  (MARS) | The app quality mean scores assessed using the mobile apps rating scale ranged from 2.43 to 4.23 (out of 5.00). | No | NA |
| Momenimovahed Z et al. 2021 | 55 | NA | NP | NS | Globally, mainly US and Europe |
| Mostafaei A et al. 2022 | 22 | JBI-Qualitative Appraisal Instrument, a score of seven and above was qualified as high quality | The score varied from 4 to 10, with 16 studies evaluate to be at low risk of bias | No | Globally, mainly in US, Canadian and European context |
| Moujaess E et al. 2020 | 88 | Not applied | NP | No | Globally, most from China and Italy (52 of 88 studies), US (13%), France (8%), UK (6%) and other countries. |
| Muls A et al. 2022 | 51 | Mixed Methods Appraisal Tool (MMAT) | Most of the studies received a  medium score, 16 studies (30%) scored four  or five stars.  Majority of papers acquired a score of 2 (15 studies) or 3 (17 studies) out of a maximum 5 score. Limitations included small samples, non-representative samples, lack of detail relating to cancer types and treatment and methodological flaws | No | Globally, 21 countries |
| Murphy A et al. 2022 | 37 | JBI critical appraisal tools, and CHEC list. Risk of bias in a study was considered high if the “yes” score  was 4; moderate if 5–6; and low risk if the score was 7 on the JBI tools | 1 study at high risk of bias, 19 at moderate risk of bias, 16 at low risk of bias | No | Globally, mainly from USA (35%) |
| Pacheco RF et al. 2021 | 9 | Cochrane Risk of Bias Table, ROBINS-I and JBI tool | The methodological quality was considered low (1 study) to moderate (1 study) for case series and low for all cross-sectional and analytical cross-sectional studies. | NS | USA, Italy, China, Spain, UK, and Iran |
| Rohilla KK et al. 2021 | 6 | Studies were assessed for validity and authenticity by five  experts from different oncology departments | NP | NS | India |
| Salehi F et ak. 2022 | 16 | NA | NP | NS | Globally, half from US (8 studies), Canada, Brazil, China, Italy, Turkey |
| Sun P et al. 2021 | 6 | NA | NP | No | Italy (2 studies), America (2 studies) England (2 studies) |
| Zapala J et al. 2022 | 160 | NA | NP | NS | NP |
| Zhang L et al. 2022 | 40 | The JBI tool | 11, 14 and 15 studies had high, unclear, and low risk of bias, respectively. Risk of bias was relatively small, indicating moderate methodological quality | No | Globally, mainly from China, Italy, Netherlands, America, and Canada |
| AlkatouI et al. 2021 | 16 | Newcastle-Ottawa Quality Assessment Form | 15 studies good quality and one article moderate quality | Yes | Globally, USA, Taiwan, Belgium, Netherlands, Japan, Italy, England, Austria and Canada |
| Cosimo SD et al. 2022 | 56 | “Risk of bias instrument for cross-sectional  surveys of attitudes and practices” from the CLARITY Group at  McMaster University | From 1 to 5, all studies (except for one) scored 3 or less, suggesting moderate to high risk of bias. | Yes | Globally, 121 countries |
| Fancellu A et al. 2022 | 7 | Not Applied | NP | Yes | Italy |
| Ferrara P et al. 2022 | 33 | Newcastle-Ottawa Scale (aNOS), reports achieving an aNOS score of 5 or greater were considered high-quality studies | Methodological quality varied across the 34 studies, of which 32 scored five or more stars on the aNOS quality assessment, while two were classified as low quality given the high risk of bias. Major reasons for bias across studies included lack of representativeness of the sampled participants, as well as substandard assessment of the outcomes as they were mostly self-reported through questionnaires. | Yes | Globally, 16 studies were carried out in America, 13 in Europe, three in Asia, and two in Africa |
| Gadsden T et al. 2022 | 17 | JBI, ROBINS-I risk of bias  tool, the Cochrane Collaboration to  assess non-randomized studies of interventions | Moderate to high risk of bias. The majority of studies outlined clear time periods for comparison, the study setting, outcomes for measurement and their data source. Comparatively, sample size was not reported in few studies, and studies commonly did not explain their statistical analysis methods nor provide confidence intervals. No study controlled for confounding. None of the included studies were scored a high methodological quality. | Yes | Majority conducted in India (13 studies), Indonesia (1 study), Sri Lanka (2 studies), Bangladesh (1 study) |
| Hesary FB et al. 2022 | 22 | The Newcastle-Ottawa Quality Assessment Form | Low to moderate risk of bias; 17 studies defined as good quality and 5 studies as fair quality | Yes | Globally, Italy, UK, Portugal, Netherlands, China, India, Japan, Turkey, Iran, Singapore |
| Lignou S et al. 2022 | 32, only one specifically to cancer, some on multiple chronic diseases | Not applied | NP | Yes, but NS | Globally, the cancer data mainly from UK |
| Majeed A et al. 2022 | 132, 60 evaluating cancer care delivery during pandemic | GRADE | Low to moderate within care delivery studies; 23 very low certainty, 12 low, 8 moderate and one high certainty. For some studies was not possible to do the assessment | Yes, but NS | Globally, Italy (6 studies), US (19 studies), France (3 studies), Poland (4 studies), Saudi Arabia (2 studies), Spain (5 studies), Greece (2 studies) and other countries. |
| Mayo M et al. 2021 | 13 | Newcastle-Ottawa Quality Assessment Tool and GRADE | Moderate to low risk of bias; 6 studies at low risk of bias, 7 at moderate risk of bias.  Based on GRADE, quality of evidence  to be high for diminished colon cancer  screening and moderate for the diminution of  breast and cervical cancer screening during  the beginning of the COVID-19 pandemic | Yes | Globally, Italy, Australia, Taiwan (3 studies), US (3 studies), France and Netherlands |
| Mazidimoradi A et al.2021 | 43 | Newcastle-Ottawa Quality Assessment Scale | Low to moderate risk of bias; 22 articles defined as good quality and 16 articles as medium quality, 5 as poor quality | Yes | Globally, most eligible studies have  been conducted in European countries 38 articles as well  as India and China. |
| Mazidimoradi A et al. 2022 | 25 | Newcastle-Ottawa Quality Assessment Scale | Low to medium risk of bias; 22 articles were defined as good quality and 3 articles as medium quality | Yes | Globally, mainly from Europe (15 studies). |
| Ng JS et al. 2022 | 31 | Newcastle-Ottawa Scale (NOS). Studies were graded by a single reviewer as high (8-9), moderate (6-7) or low (≤5) quality based on the number of stars awarded in the assessment scale | Low to moderate risk of bias; 7 studies at low risk of bias, 24 at moderate risk of bias. Most studies lost one or two points in the comparability domain for not accounting for the effects of confounders in the studies | Yes | Globally, US (8 studies), Italy (3 studies), UK (3 studies), Austria (2 studies) and Taiwan (2 studies). |
| Nikolopoulos M et al. 2022 | 15 | Newcastle–Ottawa Scale | Moderate to high risk of bias; The score ranges from 3 to 7, out of 9 maximum points. | Yes, but NS | Globally, three were conducted in the USA, two in  China and one in each of the following countries: Turkey,  Italy, Spain, India, and Austria. The online studies were conducted  in USA (two), India, and two of them globally |
| Pararas N et al. 2022 | 10 | Newcastle–Ottawa scale (NOS). Each study is awarded a score of 0 to 9 stars, quantifying its  methodological quality, with studies scoring 7 to 9 being of high quality, studies with  scores from 5 to 6 being of mediocre quality, and studies with a score of 4 or less being of  poor quality. | Low to moderate risk of bias; eight studies had an NOS score of 7 to 9  and were deemed of high methodological quality, and two studies had a score of 5 or 6 and  were deemed of mediocre methodological quality. The median score of the obtained NOS  scores were 7.5 | Yes | Globally, five studies being from east Asia (one from Japan, two from China, and two from Korea) and the remaining five from Europe (two from the United Kingdom, one from Italy, one from Ireland, and one from Serbia). |
| Riera R et al. 2021 | 62 | ROBINS-I, JBI (the studies were  categorized as presenting high quality [scored 7 or 8],  moderate quality [scored 6 or 5], or low quality [scored 4 or  lower]), IH Quality Assessment Tool for Case Series  Studies and Center for Evidence-Based Management’s critical appraisal of a survey (for survey assessment, considering the 12 questions to be answered, at the discretion of the review authors, the studies were categorized as presenting high quality [scored 9-12], moderate quality [scored 5-8], or low quality [scored 4 or lower]). | Moderate to high risk of bias; The methodological quality was considered low for case series, low for longitudinal studies, and moderate to low for cross-sectional studies. Among analytical cross-sectional studies, the quality was considered moderate for and low for the remaining 14. For surveys, the methodological quality was considered moderate for and low for the remaining eight. | Yes | Globally, majority from Italy (30.6%), USA (16.1%), China (9.7%), France (6.5%), UK (4.8%), Canada (3.2%) and the rest with 1.6% each |
| Sarich P et al. 2022 | 44 | ROBINS-I and Risk of Bias checklist for prevalence studies by Hoy Damian et al. 2012 | High risk of bias; all cross-sectional studies were at high risk of bias, the before and during studies were mix, from low to high risk of bias. The two major  sources of bias were selection of participants into the  study, mainly due to non-representative participants or  low response rates, and in the measurement of the outcome  with different methods and/or tools/questions  used before and during the pandemic. Another major source of  bias being that study populations were not representative  of the target population | Yes | Globally, across 24 countries |
| Sabeena S et al. 2022 | 7 | National Institute of Health Checklist (NHLBI, NIH).  The studies with a minimum score of eight or above,  seven, or five or less than five “Yes responses” were  considered good, fair, and poor quality, respectively. For  cross-sectional and case-control studies, question numbers  1, 2, 3, 4, 5, and 11 were applicable. The responses to the  remaining eight questions (6-10,12,13,14) were marked as  not applicable (NA). Each question was categorized as Yes,  No, others-CD (can-not determine), NA (not applicable),  NR (not reported). The studies with six “Yes” responses  were considered good, and those with four /five were taken  as fair. The studies with less than four “Yes responses”  were considered of poor quality. Two reviewers assessed  the quality of the studies. | Low risk of bias; all  these studies were qualified as good. | Yes | Globally, Slovenia, Italy, Canada, Scotland, Belgium, and US |
| Tang G et al. 2022 | 14 | Newcastle-Ottawa Scale | Low risk of bias | Yes | Globally, Turkey, China, UK, Italy, Denmark, Austria, Australia |
| Teglia F et al. 2022 | 39 | Critical  Appraisal Skills Programme for qualitative research,15 for  a maximum score of 10 points. Studies obtaining less than 7  points were considered inadequate and excluded (no article  was excluded because of a low-quality score). | Low to moderate risk of bias; all studies scored 7 or higher | Yes | Globally, America, Asia, Europe, |
| Teglia F et al. 2022 | 47 | Critical  Appraisal Skills Programme for qualitative research,15 for  a maximum score of 10 points. Studies obtaining less than 7  points were considered inadequate and excluded (no article  was excluded because of a low-quality score). | Low to moderate risk of bias; all studies scored 7 or higher | Yes | Globally |
| Thomson JD et al. 2020 | 54 | The American Society of Radiation Oncology (ASTRO) classification The ASTRO scale defines 4 levels of quality of evidence: high, moderate, low, and expert opinion. To be designated high quality, the fractionation schedule had to be supported by 2 or more well conducted and highly generalizable randomized clinical trials or meta-analyses of such trials | Mixed quality, cancer-specific results. In general, low to moderate quality. | Yes |  |
| Vigliar E et al., 2020 | 41 respondents | Not applicable | Not applicable | Yes | Globally, 23 countries |

CEBM, Critical appraisal tool of qualitative studies from Centre of Evidence-based Medicine (CEBM), University of Oxford; ASTRO, The American Society of Radiation Oncology; CASP, https://casp-uk.net/casp-tools-checklists/; CHEC, Consensus on Health Economic Criteria: CLARITY, “Risk of bias instrument for cross-sectional surveys of attitudes and practices” from the CLARITY Group at McMaster University"; JBI, Joanna Briggs Institute; MARS, Mobile Apps Rating Scale; MMAT, Mixed Methods Appraisal Tool; NHLBI, NHI, National Institute of Health Checklist; NOS, Newcastle-Ottawa Quality Assessment; NP, not provided; RBC, Risk of Bias Checklist for Prevalence Studies by Hoy Damian et al. 2012

**Appendix 6**: Summary of results on treatment modification during Covid-19 pandemic

| **Author, year of publication** | **Cancer type** | **Conclusion** |
| --- | --- | --- |
| Adham M et al. 2022 | Head and neck cancer | Medically Necessary, Time-sensitive scoring combined with Guideline from Department of Otolaryngology at Stanford University prioritizing criteria can be helpful in decision making of stratifying Risk and prioritizing surgery in head and neck cancer management. |
| Alom S et al., 2021 | Any cancer type, and per different types | Many centers judiciously considered risks and benefits for treatment continuation or initiation for patients, such as treatment-related complications and intensive care availability. Although downscaling treatment plans in cancer patients was a significant intervention in this review, there were concerns over potential undertreatment of cancer patients as a result of these treatment changes. The general consensus was that each patient should be assessed on a  case-by-case basis by multidisciplinary teams and that delaying treatments for curable cancer was not recommended. Tumor stage, histology, age, treatment type, comorbidities, patients’ general well-being and history of recent pneumonitis were taken into account when assessing the risks and benefits of  cancer treatments. Documentation of treatment variation into trust databases and regular auditing of clinical activity was also deemed crucial in maintaining standard of care during COVID-19 pandemic |
| Garg PK et al. 2020 | Any cancer type; data on 12 types of cancer | Majority guidelines for various types of cancers favored a delay in  treatment or a nonsurgical approach wherever feasible. Available guidelines are based on a low level of evidence and have significant discordance for the role and timing of surgery, especially in early tumors. |
| Gascon L et al. 2020 | Head and Neck cancer | Recommendations include adjustments regarding new patients’ referral such as performing a pre-appointment triage and working in telemedicine when possible. Surgical prioritization must be adjusted in order to respect pandemic requirements. High-grade malignancies should, however, not be delayed, due to potential serious consequences. Many head and neck interventions being aerosol-generating procedures, COVID-19 testing prior to a surgery and adequate PPE precautions are essential in operating room.  Many guidelines downsize the role of surgery in the pandemic era sometimes recommending non-surgical treatment over surgical treatment as it appears that head and neck cancer surgeries such as transoral laser or robotic surgery are aerosol-generating and as such, are  considered unsafe in this pandemic era. This needs to be weighed against the daily back and forth travel from home to the hospital for 6 to 7 weeks of patients receiving radiation therapy which might increase the risk of contamination of the patients and the staff involved in their care. |
| Hojaij FC et al. 2020 | Head and Neck, and otorhinolaryngology | Cancer should be treated, and each case should be assessed individually. Surgical prioritization must be adjusted in order to respect pandemic requirements. High-grade malignancies should, however, not be delayed, due to potential serious consequences. Tracheostomies should be performed with extreme caution and the cuff should be always insufflated below the opening area.  The use of adequate PPE (N95 mask or PAPR) with complete gown in aerosol-producing procedures should become mandatory until there is control of the epidemy, that is, vaccine or effective antiviral drugs. Endoscopic exams should only be performed if their result may change the patient’s treatment; otherwise, patient’s evaluation should be restricted to clinical exam. Evaluation of the COVID-19 status of all inpatients, especially surgery patients, should be performed to access transmission risk.  Telemedicine can help reduce hospital and outpatient visits. All the recommendations should become standard as long as there is no treatment or vaccine for the SARS-CoV-2. |
| Moujaess E et al. 2020 | Any cancer site | In the absence of universal guidelines, most of the strategies adopted involve prioritizing urgent situations such as acute leukemia, curative treatments for aggressive diseases, and adjuvant and neoadjuvant therapies while withholding or postponing palliative therapies for poor prognosis patients. Telemedicine was also encouraged. Measures to protect medical staff are proposed because this indirectly impacts patients’ safety. These measures consist of prioritizing laparoscopic procedures in cancer surgery to minimize the exposure to aerosolized specimen and limiting endoscopic diagnostic procedures to the necessary with application of strict protective measures particularly in bronchoscopy. Some medical and imaging oncology wards were completely re-organized to safely accommodate cancer patients. A detailed description of Chinese, Italian and French experience is provided, as well a summary of international guidelines for management and care of cancer patients. |
| Sun P et al. 2021 | Breast cancer reconstruction | Of the 6 included studies, 4 studies recommended the use of breast implants or tissue expansion for breast reconstruction surgery and had good results in their clinical practice. In addition, 1 study planned to use autologous free tissue transfer for breast reconstruction, and 1 study planned to use microsurgical techniques for breast reconstruction. But these 2 technologies are still in the planning stage and have not yet been implemented. |
| Cosimo SD et al. 2022 | Any cancer site, and cancer-specific including brain, head and neck, gynecological, breast, hepato-bilio-pancreatic, hematological, colorectal, skin, pediatric, urinary tract, esophagogastric, neuroendocrine, lung and soft tissues cancers. | Changes in treatment plans occurred in 65% of the centers |
| Hesary FB et al. 2022 | Gastric Cancer | Most people start complementary therapy on their own and are unaware of its side effects. Changes in the treatment of patients depend on the severity of the epidemic and risk factors in patients such as age over 75 years, comorbidities including hypertension, diabetes, liver failure, kidney, and lung failure. |
| Mazidimoradi A et al.2021 | Colorectal cancer | Changes in patients’ treatment plans and complete to partial cessation of hospitals activities—that provided treatment services—were reported. |
| Nikolopoulos M et al. 2022 | Gynecological cancer, endometrial, cervical, ovarian, and vulval cancer | - Diagnoses   Cancer surgery, chemotherapy, and radiotherapy should continue as high priority practices. gynecologic cancer surgery can be performed safely when appropriate measures for COVID-19 safety are taken. A single-center retrospective study from Madrid included 126 patients who were scheduled for surgery and suggested that with adequate preventive and protective measures, cancer surgery was possible and did not significantly compromise patients or staff |

**Appendix 7**: Summary of results on delays and/or cancellation of cancer treatment during Covid-19 pandemic

| **Author, year of publication** | **Cancer type** | **Conclusion** |
| --- | --- | --- |
| Dhada S et al. 2021 | Any cancer type | Participants in most studies reported treatment delays for a number of varied reasons including hospital cancellations, city lockdowns and COVID-19 testing requirements.  Postponement and delays in cancer screening and treatment, drug shortages and inadequate nursing care were commonly experienced by patients. Hospital closures, resource constraints, national lockdowns, and patient reluctance to use health services due to infection worries contributed to the delay. Financial and social distress, isolation, and spiritual distress were also commonly reported. Caregivers in addition felt anxious about infecting cancer patients with COVID-19. |
| Jammu AS et al. 2021 | Any cancer | Survivors have seen significant delays and cancellations of follow-up appointments and the replacement of most in person interactions with  telehealth. The COVID-19 pandemic has drastically reduced the frequency  and types of care survivors are able to access as jurisdictions attempt to reduce risk of COVID-19 transmission through in person healthcare encounters. A study showed 44% of cancer survivors reported delays across all aspects of cancer care and treatment. |
| Pacheco RF et al. 2021 | Any cancer site, including breast cancer, head and neck cancer and lung cancer. | There were delays and interruption reported on radiotherapy, surgery, treatments and outpatients’ visits. The only comparative study reported a 48.7% reduction observed in the number of outpatient visits to the hospital accompanied by a small reduction in imaging and an improvement in radiation treatments after the implementation  of a multiple organizational strategy. |
| Zapala J et al. 2022 | Any cancer site | For 12 weeks after the COVID-19 outbreak in Europe, 72.3% of all scheduled operations were cancelled. Oncological procedures were the second most numerous groups among them: 37.7% of them were postponed (i.e., 2,324,070 out of 6,162,311). In Poland, 22,656 surgeries were cancelled during the period indicated (COVIDSurg Collaborative, 2020). In Poland, since the announcement of the pandemic and lockdown, the number of issued DiLO Diagnostic and Oncological Treatment cards has decreased. Data from the National Health Fund (NFZ) show that already in the first month of the epidemic in Poland (March 2020) there were 1780 fewer of them than a year earlier |
| Cosimo SD et al. 2022 | Any cancer site, and cancer-specific including brain, head and neck, gynecological, breast, hepato-bilio-pancreatic, hematological, colorectal, skin, pediatric, urinary tract, esophagogastric, neuroendocrine, lung and soft tissues cancers. | Cancellation/delay of treatment occurred in 58% of centers; delay of outpatient visits in 75%; changes in treatment plans in 65%; and a general reduction in clinical activity in 58%. |
| Ferrara P et al. 2022 | Cervical cancer | All but one study that investigated cervical cancer treatment reported changes in the number of women with cervical lesions who received treatments, as well as treatment delay and interruption. With a major impact during the first wave in 2020, COVID-19 and restriction measures resulted in a substantial disruption in cervical cancer prevention and management, with declines in screening and delays in treatment. |
| Gadsden T et al. 2022 | Any cancer site and cancer-specific, including cervical cancer (n=1), paediatric (n=1), oral (n=1), blood (n=1), gastrointestinal (n=1) and head and neck (n=1) cancers | Compared to a pre-pandemic period, 10/17 cancer studies found a >40% reduction in outpatient services. Seven studies reported on inpatient admissions finding reductions ranging from 14.4 to 61.6%. To some extent, the magnitude of service reduction reported, depended on the timeline of the study. Studies that only analysed service provision during a lockdown period were likely to report higher reductions than those that covered the whole of 2020. Compared to same period in 2019, the number of outpatient services reduced by 8% (p=0.002, 95% CI: 6.2 to 9.5%) during the pandemic period, while inpatient admissions decreased by 26% (p=0.002, 95% CI: 22.9 to 30.3%).  Of these 17 studies, only two reported on the impact of mitigation measures to maintain service provision during the pandemic. In the absence of any national guidelines in India, Mallick and colleagues prioritized radiotherapy treatment for oncology patients, continued services for patients already undergoing treatment and deferred new starts for adjuvant therapy. Additionally, a staff rotation policy was implemented to ensure that human resources could be redeployed to prevent delays and deliver full services for those with the highest priority. Although outpatient consultations dropped by 58% during lockdown, more than 90% of high-priority cancer treatments (specifically radiotherapy and chemotherapy) were implemented as planned. |
| Lignou S et al. 2022 | Pediatric cancer | After recommendations by professional bodies and commissioners, multiple changes to cancer care have been established since the start of the pandemic, from the point of diagnosis (e.g., suspension of screening services) to treatment plans.  Between the 18th and 31st January 2021, pediatric and noncancer elective surgical activity was occurring at less than a third of the rate of the previous year. |
| Majeed A et al. 2022 | Pediatric cancer | The pandemic has resulted in delays and interruptions to cancer therapies and delays in childhood cancer diagnoses in both HICs and LMICs. However, these ﬁndings were disproportionately reported in LMICs, with signiﬁcant staff shortages, supply chain disruptions, and limited access to cancer therapies for patients. |
| Mazidimoradi A et al.2021 | Colorectal cancer | Treatment of colorectal cancer has also decreased significantly or has been delayed, interrupted, or stopped. This reduction and delay have been observed in all treatments, including surgery, chemotherapy, and long-term radiation therapy; only cases of emergency surgery and short-term radiotherapy has increased. The waiting time for hospitalization and the length of hospital stay after surgery has been reported to be higher. Changes in patients’ treatment plans and complete to partial cessation of hospitals activities—that provided treatment services—were reported. |
| Nikolopoulos M et al. 2022 | Gynecological cancer, endometrial, cervical, ovarian, and vulval cancer | Severe delays in management have been reported. The percentage of the patients experiencing delay in treatment is consistently more than 10% across the studies identified with most of them being in surgical treatment. There is also a move towards conservative management, with hormonal treatment being utilized in the treatment of endometrial cancer and neoadjuvant chemoradiotherapy being performed in cases, which would be treated with primary surgery before the pandemic. Surgical management has changed with an increased rate of laparotomies compared to laparoscopies despite the preventive measure taken. |
| Riera R et al. 2021 | Any cancer site, and cancer specific including breast, head and neck, urological, colorectal, skin, hematological, gynecological, pediatric, lung, hepatopancreatobiliary, stomach and musculoskeletal. | Frequent determinants for disruptions were provider- or system-related, mainly because of the reduction in service availability. The studies identified 38 different categories of delays and disruptions with impact on treatment, diagnosis, or general health service. Delays or disruptions most investigated included reduction in routine activity of cancer services and number of cancer surgeries; delay in radiotherapy; and delay, reschedule, or cancellation of outpatient visits. Interruptions and disruptions largely affected facilities (up to 77.5%), supply chain (up to 79%), and personnel availability (up to 60%). |
| Teglia F et al. 2022 | Any cancer site and cancer-specific | An overall reduction of −18.7% (95% CI, −24.1 to −13.3) in the total number of cancer treatments administered during the COVID-19 pandemic compared to the previous periods. Surgical treatment had a larger decrease compared to medical treatment (−33.9% versus −12.6%). For all three types of treatments, we identified a U-shaped temporal trend during the entire period January–October 2020. Significant decreases were also identified for different types of cancer, in particular for skin cancer (−34.7% [95% CI, −46.8 to −22.5]) and for all geographic areas, in particular, Asia (−42.1% [95% CI, −49.6 to −34.7]). Conclusions and Relevance: The interruption, delay, and modifications to cancer treatment due to the COVID-19 pandemic are expected to alter the quality of care and patient outcomes. |

**Appendix 8**: Summary of results on delays and/or cancellation of cancer screening during Covid-19 pandemic

| **Author, year of publication** | **Cancer type** | **Conclusion** |
| --- | --- | --- |
| Dhada S et al. 2021 | Any cancer type | Disruption to cancer screening and diagnosis was a commonly reported theme. A cross-sectional survey assessing the experiences of sarcoma patients at two of the largest specialist sarcoma centres in Europe reported that one-third experienced postponement of appointments or scans by at least three months. Patients also reported cancellations of routine follow-up clinic appointments in studies conducted in the UK and US.31,34 In a further two studies, patients expressed anxiety and fear due to postponement of cancer-related laboratory tests and diagnostic imaging. |
| AlkatouI et al. 2021 | Any cancer site, breast cancer, colorectal cancer, gynecological cancer, lung cancer, colon cancer | - The impact of COVID-19 was categorized into four dimensions: a significant decline in cancer screening and pathology samples, the cancer diagnosis rate, an increase in advanced cancers, mortality rate and years of life lost (YLLs). Published studies have disclosed a marked decline in cancer screening, diagnostic imaging, as well as histopathological and cytological biopsies during the COVID- 19 pandemic. The effect was more pronounced in countries with a greater prevalence of COVID-19 or poorly controlled rates of COVID-19 infection. - Compared to the pre-COVID period, colonoscopy rates fell by 4.1% to 75%. Gastroscopies, prostate, and lung screening rates were reduced by 57%, 74%, and 56%, respectively. Screening mammograms declined by 22.2-85%. - The histopathological and cytological workload was reduced by 35-72% compared to the preceding three years. Reductions in cancer biopsies were reported for breast (−31 to -71%), colon (-33 to -79%), and lung cancer (-47 to -58%). |
| Fancellu A et al. 2022 | Colorectal Cancer | - We found that reduction of CRC screening activity surpassed 50% in most endoscopic units, with almost 600000 fewer CRC screening exams conducted in the first 5 months of 2020 vs the same period of 2019. While the consequences of the discontinuation of endoscopy screening for the prognosis and mortality of CRC will be evident in the next few years, recent data confirm that CRC is currently treated at a more advanced stage than in the pre-COVID-19 era. |
| Ferrara P et al. 2022 | Cervical cancer | - Reports on cervical screening and cancer diagnosis activities showed a substantial impact of the pandemic on access to screening services and diagnostic procedures. With a major impact during the first wave in 2020, COVID-19 and restriction measures resulted in a substantial disruption in cervical cancer prevention and management, with declines in screening and delays in treatment. |
| Hesary FB et al. 2022 | Gastric Cancer | - The COVID-19 epidemic has reduced the number of screenings, In Italy, the number of endoscopies has decreased by 53.6% compared to 2019 . In the Netherlands, the rate of gastroscopy has decreased by 57%. The number of endoscopies depends on the duration and severity of the restrictions associated with COVID-19 |
| Mayo M et al. 2021 | Any cancer site, but identified mainly   - breast   cancer   - colon cancer - cervical cancer - lung cancer | - Incidence rate ratios were significantly lower for screening during the COVID-19 pandemic for breast cancer (0.63; 95% CI, 0.53 to 0.77; P<.001), colon cancer (0.11; 95% CI, 0.05 to0.24; P<.001), and cervical cancer (0.10; 95% CI, 0.04 to 0.24; P<.001). |
| Mazidimoradi A et al. 2022 | Colorectal cancer | Screening has decreased from 28 to 100% in different countries and at different times after the onset of the COVID-19 pandemic. During this period, only 2 to 2.5% of hospitals and screening centers with 100% capacity continued to operate, and more than 77% of them limited their activities to less than 10% of their normal capacity. Also, completion of colonoscopies requiring examination showed a decrease of 65.7%, surveillance colonoscopy showed a decrease of 44.6 to 79%, prescription colonoscopy decreased 60 to 81%, and referrals to colonoscopy showed a 43% decline. However, emergency colonoscopy shows a 2 to 9% increase. The use of the Fecal immunochemical test (FIT) test is also generally declining but is increasing in areas used as a colonoscopy alternative |
| Ng JS et al. 2022 | Breast cancer | Cancer screening rate dropped by an estimated 41–53% between 2019 and 2020. No differences in mammogram screening rates depending on patient age or ethnicity were observed. However, countries that implemented lockdown measures were associated with a significantly greater reduction in mammogram and diagnosis rates between 2019 and 2020 in comparison to those that did not. |
| Sabeena S et al. 2022 | Cervical Cancer | The pooled proportion of women screened for cervical cancer in 2019 was 9.79% (95% CI 6.00%-13.59%, 95% prediction interval 0.42%-23.81%). During the pandemic, the pooled proportion of screened women declined to 4.24% (95% CI 2.77%-5.71%, 95% prediction interval 0.9%-17.49%). |
| Teglia F et al. 2022 | Breast cancer, colorectal cancer and cervical cancer | There was an overall decrease of −46.7% (95%CI, −55.5%to −37.8%) for breast cancer screening, −44.9% (95%CI, −53.8% to −36.1%) for colorectal cancer screening, and −51.8%(95%CI, −64.7%to −38.9%) for cervical cancer screening during the pandemic. For all 3 cancers, a U-shaped temporal trend was identified; for colorectal cancer, a significant decrease was still apparent after May 2020 (in June to October, the decrease was −23.4%[95%CI, −44.4%to −2.4%]). Differences by geographic area and screening setting were also identified. |
| Vigliar E et al., 2020 | Any cancer, Cytopathology Practice | The COVID-19 pandemic resulted in a drastic reduction in the total number of cytology specimens regardless of anatomic site or specimen type. The rate of malignancy increased, reflecting the prioritization of patients with cancer who were considered to be at high risk. Overall, the sample volume was lower compared with 2019 (104,319 samples vs 190,225 samples), with an average volume reduction of 45.3% (range, 0.1%-98.0%). The percentage of samples from the cervicovaginal tract, thyroid, and anorectal region was significantly reduced (P < .05). Conversely, the percentage of samples from the urinary tract, serous cavities, breast, lymph nodes, respiratory tract, salivary glands, central nervous system, gastrointestinal tract, pancreas, liver, and biliary tract increased (P < .05). An overall increase of 5.56% (95% CI, 3.77%- 7.35%) in the malignancy rate in nongynecological samples during the COVID-19 pandemic was observed. When the suspicious category was included, the overall increase was 6.95% (95% CI, 4.63%-9.27%). |

**Appendix 9**: Summary of results on delays and/or reduced cancer diagnoses during Covid-19 pandemic

| **Author, year of publication** | **Cancer type** | **Conclusion** |
| --- | --- | --- |
| AlkatouI et al. 2021 | Any cancer site, breast cancer, colorectal cancer, gynecological cancer, lung cancer, colon cancer | - There was a decline in cancer diagnosis rate, which varied by type of cancer. In Italy, there was a 11% decline in cancer diagnosis. In Canada, a three- and six-month interruption in the breast screening program caused a 7% and 14% drop in cancer diagnosis. A marked drop in newly diagnosed gynecological tumors (-24% to -49%) was reported in Austria during theCOVID-19 pandemic, and the median age of the patients was significantly lower than that of patients diagnosed with cancer in 2019 (59.4 vs. 61.3 years). A nearly 10% decline was noted in the diagnosis of breast cancer. - The interruption of cancer prevention due to suspended cancer screenings may delay the diagnosis, increase the numbers of symptomatic patients, and disclose cancers in more advanced stages. According to predictions, once the lockdown has been lifted it will take a minimum of 12-24 weeks to clear the queue of missed cancer screenings - The effect of delayed cancer diagnosis will not be perceived in the immediate future alone; premature deaths may occur as long as ﬁve years later. Delayed cancer screening is estimated to cause the following additional numbers of cancer deaths   secondary to breast, esophageal, lung, and colorectal cancer, respectively: 54,112–65,756, 31,556–32,644, 86,214–95,195, and 143,081–155,238 in the worldwide   - The reduction has been attributed to stay-at-home orders, patients’ fear of infection, hesitation to seek care, the perceived risk of exposure to COVID-19 for clinicians, changing hospital policies in redeployment of staff towards critical care for the management of COVID-19 patients, triage of patients with COVID-19 infection, and the cessation of cancer screening in hospitals |
| Fancellu A et al. 2022 | Colorectal Cancer | - Recent data confirm that CRC is currently treated at a more advanced stage than in the pre-COVID-19 era. Studies reported that CRC new diagnoses decreased up to 62% in 2020 compared to previous years, while on average to 11.9% with Northern Italy experiencing the highest decrease |
| Ferrara P et al. 2022 | Cervical cancer | - Eight studies specifically analyzed the impact of COVID-19 on cervical cancer diagnosis and diagnostic procedures, comparing 2020 with the pre-pandemic period. - 25.7% less cancer cases between May and October 2020 in United Kingdom (Davies et al., 2022) - -7% in April–June 2020 in South Africa, and -73.4% during the whole 2020 year in Portugal - A decrease of 13% in diagnostic invasive procedures in Slovenia in 2020 - In Brazil Bonadio et al. patients had a more advanced-stage at diagnosis during the with the proportion of stages III-IVA increased by 13.5% |
| Hesary FB et al. 2022 | Gastric Cancer | - The average gastric cancer diagnosed per week decreases by 54.1%. During the COVID-19, inappropriate endoscopic referrals decreased. Although the number of endoscopies has decreased, the incidence of   gastric cancer has increased significantly. In other words, the  rate of gastric cancer diagnosed has increased in endoscopies |
| Lignou S et al. 2022 | Pediatric cancer | - After recommendations by professional bodies and commissioners, multiple changes to cancer care have been established since the start of the pandemic, from the point of diagnosis (e.g., suspension of screening services) to treatment plans. - Data collected from Salford in the UK found a large decrease in the rate of new diagnoses for circulatory system diseases, type 2 diabetes, malignant cancers and common mental health problems. Another UK-based study supported these results for cancer patients. Screening services were suspended and there was an 80% decrease in 2-week wait cancer referrals since March 2020 due to reduced diagnostic services including endoscopies, social distancing rules (including instructions for the public to present at GPs with urgent concerns only) and public health anxiety. - The use of telehealth for people with cancer suggests a greater proportion of missed diagnoses. - A study on the impact of delays in cancer diagnosis in adults and children estimated that between 3291 and 3621 avoidable deaths will have occurred from 5 cancer types in the 5 years after diagnosis compared with the pre-pandemic period. An additional 59,204–63,229 years of life lost will be attributable to delays in cancer diagnosis alone as a result of the ﬁrst COVID-19 lockdown in the UK. |
| Majeed A et al. 2022 | Pediatric cancer | - Altogether, many HICs reported less pediatric oncology diagnoses when compared to before the pandemic, especially in relation to solid tumor diagnoses |
| Mazidimoradi A et al.2021 | Colorectal cancer | - A delay in the diagnosis of colorectal cancer has reported from 5.4 to 26% - Decreased diagnosis of new cases of colorectal cancer during COVID-19 pandemic is seen in most countries, as Spain has reported 48% reduction and Brazil has reported 46.3% reduction in the diagnosis of colorectal cancer - Most studies have reported that the diagnosis of colorectal cancer has been significantly reduced. This reduction included a reduction in the overall diagnosis of cancer, a reduction in routine referrals, and a reduction in cancer detection through screening programs. At least 2828 colorectal cancer cases have not been diagnosed in the UK. |
| Ng JS et al. 2022 | Breast cancer | - Cancer diagnosis rates dropped by an estimated 18–29% between 2019 and 2020. However, countries that implemented lockdown measures were associated with a significantly greater reduction in mammogram and diagnosis rates between 2019 and 2020 in comparison to those that did not. |
| Nikolopoulos M et al. 2022 | Gynecological cancer, endometrial, cervical, ovarian, and vulval cancer | - The number of new diagnoses has declined |
| Vigliar E et al., 2020 | Any cancer, Cytopathology Practice | - The COVID-19 pandemic resulted in a drastic reduction in the total number of cytology specimens regardless of anatomic site or specimen type. The rate of malignancy increased, reflecting the prioritization of patients with cancer who were considered to be at high risk. An overall increase of 5.56% (95% CI, 3.77%- 7.35%) in the malignancy rate in nongynecological samples during the COVID-19 pandemic was observed. When the suspicious category was included, the overall increase was 6.95% (95% CI, 4.63%-9.27%). |

**Appendix 10**: Summary of results on psychological distress of cancer patients during Covid-19 pandemic

| **Author, year of publication** | **Cancer type** | **Conclusion** |
| --- | --- | --- |
| Ayubi E et al. 2021 | Any cancer type | Overall prevalence of depression and anxiety were 0.37 (0.27, 0.47); I2 = 99.05%, P value < 0.001 and 0.38 (0.31, 0.46); I2 = 99.08%, P value < 0.001, respectively. Compared to controls, cancer patients had higher anxiety level [standard mean difference (SMD 0.25 (95% CI 0.08, 0.42)].  The findings of this study suggest that the prevalence of depression and anxiety among patients with cancer during the COVID-19 pandemic can reach considerable levels, although observed substantial heterogeneity should be considered when interpreting the results. |
| Dhada S et al. 2021 | Any cancer type | Patients and caregivers experienced delays in cancer screening, treatment and  care during the COVID-19 pandemic and negatively affected their psychological wellbeing. Major recurring themes of barriers to accessing cancer screening and diagnosis were identified; anxiety and fear; perceived risks of infection; adverse impact on personal life, family, and finances; caregivers concerns and resilience; and coping mechanisms adopted by patients and carers.  Feelings of anxiety and fear surrounding the pandemic were common, with eighteen studies referencing changes in emotional and psychological functioning. Notably, cancer patients expressed fear about the consequences and complications arising from them contracting COVID-19.17 There were emotional reactions to the prospect of not being able to say a final farewell to family and friends. References were also made to fears surrounding family members contracting the virus and patients expressed worries and concerns about treatment delays due to the postponement of elective procedures. More than half of the participants in a US study reported new onset of anxiety or depression. Nearly a quarter (23% of the 204) participants in one study were in receipt of the government “shielding” advice.  Studies conducted in Saudi Arabia and Iran examined anxiety responses amongst children and their caregivers. Parents reported fears surrounding COVID-19 mortality rates and expressed concerns about the high transmissibility and limited knowledge surround. In one of these studies, over two-thirds of parents reported the onset of new behavioral issues amongst their children since the pandemic.17 Parents were worried about the negative effects of the pandemic on children’s mental and physical health, both now and in the long-term. |
| Jammu AS et al. 2021 | Any cancer | The COVID- 19 pandemic may detrimentally impact the psychosocial and  physical wellbeing of cancer survivors. Available articles also highlight that increased anxiety and distress has been observed among some survivors with respect to accessing telehealth services and doubts related to the quality  of care available through the remote medium. Strict social distancing restrictions across jurisdictions have also reduced access to social and support networks that some survivors may have previously depended upon for physical and psychosocial support. Preliminary research also indicates  that cancer survivors are more prone to catastrophizing and health anxiety in relation to COVID-19 compared to healthy controls. |
| Kirby A et al. 2022 | Any cancer type, providing data for 7 cancer-specific sites | The pandemic exasperated existing psychological strain and associated adverse outcomes including worry and fear (of COVID-19 and cancer prognosis); distress, anxiety, and depression; social isolation and loneliness. |
| Legge H et al. 2022 | Any cancer type, 4 cancer-specific information | The findings identified that individuals affected by cancer reported a range of  physical, psychological, social, and health system unmet needs during the global pandemic. Unique to the pandemic itself, there was fear of the unknown of the longer-term impact that the pandemic would have on treatment outcomes, cancer care follow-up, and clinical service delays. Many individuals living with cancer experienced unmet needs and distress throughout the different waves of the COVID-19 pandemic, irrespective of cancer type, stage, and demographic factors. |
| Momenimovahed Z et al. 2021 | Any cancer site, and cancer-specific information | COVID-19 greatly affects psychological health of  cancer patients. Fear of COVID-19, fear of disease progression, disruption of oncology services, cancer stage, and immunocompromised status were the most common causes of psychological distress in oncology patients which can influence patients’ decisions about treatment. Although psychological distress affects many people, it can confuse cancer patients to the point that they refuse to continue treatment for the fear of infection and worsening of their condition. |
| Muls A et al. 2022 | Various cancer types (52%, 27 studies), 48 cancer specific. | Four themes were identified: Emotional aspects  and Quality of Life; Psychosocial aspects; Impact of COVID-19 on self; Impact of COVID-19 on cancer, with  themes overlapping.  Most studies reported increased levels of anxiety and depression, but comparators were not always clear or comparable across the studies  Quality of life was reported as having been impacted negatively due to the COVID-19 pandemic.  The most common psychological aspects assessed were employment, financial difficulties, loneliness and isolation, social support, and uncertainty about the future.  Although fear of contracting COVID-19 was a big factor that increased cancer patients’ levels of anxiety, depression and distress, there are several studies reporting higher levels of anxiety and depression associated with fear of cancer recurring or progressing due to treatment delays or cancellations |
| Rohilla KK et al. 2021 | Any cancer site | During this COVID-19 pandemic phase, cancer patients have suffered from emotional trauma. Research evidence shows that the diagnosis of cancer has a strong association with a profound psychological effect, which changes a patient’s life completely and causes a persistent threat to their survival. the overall quality of life of each cancer patient is very much affected. To date, as per literature research, no study has been conducted in India to assess the quality of life of cancer patients |
| Zapala J et al. 2022 | Any cancer site | - The COVID-19 pandemic resulted in the appearance of new problems in the population of oncological patient, or it made the existing problems more severe. Therefore, it made it significantly more difficult to meet their needs on various levels and sometimes it even made it impossible. - Confronting a stressor such as the COVID-19 pandemic outbreak is associated with a reduction in psychological resilience on an unprecedented scale and with consequences that are difficult to assess. According to psychological research on the effects of disasters, an increased incidence of post-traumatic stress disorder (PTSD) and anxiety and depression syndromes is expected - The number of medical certificates for self-reported illness from the category ``Mental and behavioural disorders'' has increased significantly. Compared to 2019, the number of certificates issued increased by 25.3% and the number of days of sickness absence increased by 36.9%. - Compared to 2019, there was an increase of 21.3% in the number of certificates issued for depression and 30.4% in the number of days of absence. The number of medical certificates for depression accounted for 26.5% of certificates issued for mental and behavioral disorders and 1.9% of all certificates issued for self-reported illness in 2020. Nearly half (44.7%) of medical certificates for depression were for people aged 35-49 years - The anxiety and fear resulting from the pandemic situation are compounded by the stress of cancer - During the COVID-19 pandemic period, mean stress, anxiety and depression scores in all countries were higher than normative data except Vietnam |
| Zhang L et al. 2022 | Any cancer | - Pooled results showed that the PR of clinically significant depression, anxiety, PTSD, distress, insomnia, and fear of cancer progression/recurrence among cancer patients were 32.5%, 31.3%, 28.2%, 53.9%, 23.2%, and 67.4%, respectively. Subgroup analysis revealed that patients with head and neck cancer had the highest PR of clinically significant depression (74.6%) and anxiety (92.3%) symptoms. Stratified analysis revealed that patients with higher education levels had higher levels of clinically significant depression (37.2%). A higher level of clinically significant PTSD was observed in employed patients (47.4%) or female with cancer (27.9%). |
| Hesary FB et al. 2022 | Gastric Cancer | - The prevalence of anxiety in resistant to treatment patients and patients with advanced cancer was higher than other patients. Also, the prevalence of anxiety was higher in patients over 60 years of age - Restrictions during the epidemic have had a negative impact on people’s emotion psychologically and physiologically. - Anxiety in patients with cancer was higher than other patients. The anxiety and stress in patients with gastric cancer during corona virus outbreak increased compared to prior to corona virus. Patients’ stress caused problems such as depression, sleep disorders, cognitive impairment, and a pain. Also, patients with anxiety have worse scores on resilience, social isolation, and higher overall stress and lower performance scores. The length of time a cancer has been diagnosed and the incidence of pain have affected stress so much that the less time has passed since a person was diagnosed with cancer, the more stress they will have [26]. Patients’ anxiety varies according to the stage of the disease, oral chemotherapy or non-use of chemotherapy, age over 60 years, advanced incurable cancer, and knowledge of the purpose of treatment - Feeling of vulnerability and fear of COVID-19 have been other problems of patients. Feelings of vulnerability were associated with variables such as female gender, chemotherapy, and age over 65 years, and remained stable in 42 cases. Feelings of confusion and confusion itself, sadness and discouragement, sleep problems, lack of interest, and pleasure as well as pessimism have been other problems reported by patients. |
| Nikolopoulos M et al. 2022 | Gynecological cancer, endometrial, cervical, ovarian, and vulval cancer | - The online patient survey by Frey et al., estimated the effect that the COVID-19 pandemic had in the management and the quality of life of patients with diagnosis of ovarian cancer, with 89% of patients suffering significant anxiety, which is significantly greater that previously reported (57.9%). The main concern of the patients was reported as acquiring COVID-19 infection, followed by cancer recurrence, safety of family members, access to healthcare and financial implications |

**Appendix 11**: Summary of results on telemedicine in cancer care during Covid-19 pandemic

| **Author, year of publication** | **Cancer type** | **Conclusion** |
| --- | --- | --- |
| Alom S et al., 2021 | Any cancer type, and per different types | In summary, the COVID-19 pandemic has emphasized the benefits of the clinical application of tele-oncology for cancer patients. As many cancer treatments often result in immunosuppression, cancer patients are a prominent risk group in the pandemic due to their increased susceptibility to contracting COVID-19. Thus, a significant benefit identified for tele-oncology is that it reduces the risk of infection through decreasing in-person contact, while maintaining care continuity. Additionally, since previous literature has reported that virtual oncology services are efficient, cost effective and result in good patient satisfaction, the future of tele-oncology is a promising prospect and is likely to be continually adopted post-pandemic in routine clinical care. |
| Dhada S et al. 2021 | Any cancer type | Patients identified positive aspects of telecommunication for example, the ease of accessing care from the privacy and comfort of one’s home and the ability to maintain physical distance. Most patients in one study hoped for continuation of online services post-pandemic. In another study, almost two-thirds of patients felt that they were still able to contact the healthcare team and so expressed feelings of reassurance. However, in one study conducted in India, patients reported difficulties in booking virtual appointments and unpredictable network issues  Across the included studies, many expressed concerns with the transition to virtual appointments with their health care team, noting that the previous model of in-person appointments provided reassurance and feelings of comfort |
| Hojaij FC et al. 2020 | Head and Neck, and otorhinolaryngology | Fifteen articles from the 35 selected recommended the use of  telemedicine for triage, routine consults, or patient revaluation. Four articles recommended the use of telemedicine but did not specify the situations to be used. |
| Lu DJ et al. 2021 | Any cancer site, and cancer specific  Most apps (30/41, 73%) that met inclusion criteria were  general health/pain symptom trackers, and 11 of 41 (27%) were cancer-specific apps. Of the cancer-specific apps, 5  of 11 (46%) were nonspecific, whereas the remaining 6  included 1 each (1/11, 9%) for blood, lymphoma, head  and neck, breast, pancreatic, and ovarian cancers,  respectively. | Various symptom tracking apps are available in the mobile health  market, but the number of apps targeted toward patients with  cancer remains limited. Most apps (73%, n Z 30) were general health/pain symptom trackers, and 27% (n Z 11) were cancer specific. Only 1 app has been trialed for usability among patients with cancer |
| Mostafaei A et al. 2022 | Any cancer type, and 5 cancer-specific | Telemedicine assists but cannot be a substitute for face-to-face appointments in a health care crisis and in the provision of routine care to stable patients with cancer.”  Infrastructural drivers and healthcare provider’s support and attention affect patients’ experiences and feelings about telemedicine patients who use telemedicine expect their health care providers to devote enough time and consider emotional needs, the lack of which can develop negative response |
| Murphy A et al. 2022 | Any cancer site, multiple/all cancer types (67%). | - Telehealth adoption seemed to “bolster” the delivery of healthcare services, providing a level of continuity of care during this highly uncertain time. However, it is evidenced that a “one size fits all” approach to telehealth is not appropriate to support the delivery of essential healthcare services for cancer patients - From an economic perspective, there were costs and benefits associated with the adoption of telehealth. During the early phase of the pandemic, for some service providers, telehealth adoption consisted of using existing available technologies such as telephone consultations and video calls. This was the case for approximately 61% of the studies. For the remainder, there are costs associated with setup and maintenance of telehealth technology, including data protection. Following the early phase, strategic investments were made in hardware and software solutions to support the delivery of telehealth beyond the initial emergency response - The availability of telehealth services minimized the need for emergency admissions to acute services - Telehealth provides beneficial spillovers for patients and their families. These include reduced travel time and efficiency gains with reduced waiting times from performing consultations from home with a loved one present. However, there are also costs and access barriers, including network issues and technology costs, which disproportionately impact vulnerable groups. - From a psychological perspective, the switch to telehealth for some was associated with higher levels of cancer worry and feelings of isolation, whereby the lack of in-person access created mental health strain for patients and survivors who were forced to solely rely on telehealth communication. For some patients, this outweighs the comfort and support benefits of being at home. |
| Salehi F et ak. 2022 | Any cancer site | - The results indicated that most of the patients contacted by telemedicine services mostly used to interact with patients breast cancer (n=4, 25%). The most common use of telemedicine was the provision of virtual visit services (n=10, 62.25%). Besides, communication was most frequently provided by live video conferences (n=11, 68.75%). - Telemedicine can provide continued access to necessary health services in oncology care and serve as an important role in pandemic planning and response. |
| Zapala J et al. 2022 | Any cancer site | - In Poland during the epidemic period, 80% of medical consultations took place via telemedicine services. According to the Government Report ``Survey of Satisfaction of Patients Using Telemedicine Services with their Primary Care Physician during the COVID- 19 Epidemic Period'', almost 92% of the respondents felt that obtaining a telemedicine service helped solve their health problem (National Health Fund of Poland, 2020). 43.2% expressed the belief that telemedicine service/ video advice should be one of the main channels of contact with the primary care physician (PCP) and it should be the physician who decides whether it is necessary for the patient to visit the health facility. A total of 30.4% of respondents thought that tele-counselling was appropriate when consulting chronic, previously known health problems and continuing treatment. A total of 36.3% of respondents rated the quality of an in-patient visit higher than a teleportation; according to this group, direct contact with the doctor and the opportunity to ask questions about treatment recommendations are important. A total of 6% of respondents reported that during the telemedicine - Although telemedicine has gained acceptance by both the majority of patients and medical staff, its potential risks are also noted. Moreover, telephone communication about a patient undergoing isolation in unstable or severe clinical conditions is a difficult task for doctors, nurses and family members due to the emotional burden. |
| Cosimo SD et al. 2022 | Any cancer site, and cancer-specific including brain, head and neck, gynecological, breast, hepato-bilio-pancreatic, hematological, colorectal, skin, pediatric, urinary tract, esophagogastric, neuroendocrine, lung and soft tissues cancers. | - Virtual visits were implemented by the majority (72%) of center |
| Lignou S et al. 2022 | Pediatric cancer | - The use of telehealth for people with cancer suggests a greater proportion of missed diagnoses. |

**Appendix 12**: Summary of results on financial distress and social isolation of cancer patients during Covid-19 pandemic

| **Author, year of publication** | **Cancer type** | **Conclusion** |
| --- | --- | --- |
| Dhada S et al. 2021 | Any cancer type | Financial and social distress, isolation, and spiritual distress were also commonly reported. Impact on social activities due to lockdowns was also described, with lone lines fueling patients’ worries about their cancer. Concerns surrounding loss of income and employment instability for cancer patients and family members were reported. Those receiving palliative care expressed frustration and fear at the possibility of not being able to fulfil their last wishes. |
| Jammu AS et al. 2021 | Any cancer | The pandemic has left survivors dealing with the consequences of rigorous cancer treatment in the context of new challenges related to social isolation, financial hardship and uncertainty with respect to their ongoing care.  The pandemic and its economic consequences may disproportionately impact cancer survivors and their overall health-related quality of life and mortality. Evidence indicates that cancer survivors may be more economically vulnerable  and face greater financial hardship during the COVID-19 pandemic. |
| Kirby A et al. 2022 | Any cancer type, providing data for 7 cancer-specific sites | The economic burden associated with cancer for patients during the pandemic included direct and indirect costs with both objective (i.e., financial burden) and subjective elements (financial distress).  Several studies reported social isolation and loneliness amongst cancer patients which was mediated by detachment from loved ones, lack of social interaction, loneliness, fear of infection, worries about the future, and economic difficulties. Several at-risk groups identified as feeling isolated and anxious. These included American women with ovarian cancer (10%) and young cancer patients (aged 18–39) undergoing cancer care who felt more isolated (52%) than pre-pandemic, specifically missing social interactions. Studies reported that lower levels of education and living without minor children in Israel were associated with feelings of loneliness and isolation. In the USA cancer patients experiencing social distancing and living alone had feelings of loneliness, while disruptions to cancer care were associated with increased loneliness and social isolation. Some patients’ perceived risk of COVID-19 infection caused them to engage in extreme levels of social isolation where they had no visitors and lived alone. For others, the pandemic exasperated underlying situations. For example, upon comparing a more stressed group to a less stressed group found the former reported significantly higher levels of loneliness and social.  From an individual perspective where treatments are not covered under government schemes or insurance, patients experience a large financial burden, in-terms of out of pocket expenses, expensive health insurance premiums, or deductibles. Transition to telemedicine saved US patients’ time and money ($170). Globally, testing for COVID-19 (negative results were required to attend appointments) introduced varying fee structures with additional costs experienced amongst cancer patients in India and South Korea. These costs affected patients’ decision to undergo cancer treatment particularly in poorer countries.  Cancer patients in India (22.2%) expressed fears around losing their jobs and the implications of the expected economic crisis for their family. Similarly, in the USA, cancer patients reported concerns with maintaining employment.  These economic conditions exasperated pre-existing cancer financial burdens, including paying for prescriptions and high insurance deductibles in private healthcare markets in the USA. Patients that were unemployed, with low educational attainment, or with lower incomes, or were under financial pressure had greater difficulties accessing care in India. In the USA, non- Hispanic white, married, more educated, and older cancer patients were less likely to cite financial worries. These socio-economic inequalities coupled with increased financial burden create barriers to accessing care |
| Legge H et al. 2022 | Any cancer type, 4 cancer-specific information | People affected by head and neck cancers noted periods of social isolation, which were exacerbated at holiday times, when they could not have family or friends over to visit due to the increasing infection risk.  Patients sacrificed their self-care due to work demands [36]. Few participated in exercise or health self-management behaviors. Practically, participants expressed varying issues with transport, resulting in deficits in the accessibility of care or medications, and causing an overall disruption to their daily lives. Many experienced financial toxicities due to reduced household income. Many participants expressed concerns about reduced household income from not being able to attend work due to enforced lockdowns. Others alluded to issues with medical insurance or reimbursement, loss of employment, financial difficulties, reduction to working hours, or being forced to take sick or take annual leave. Many participants reported distressed about not being able to engage in usual activities of living with a loss of work as an additional practical concern for them. Financial concerns were a significant predictor of psychological distress with a positive association between loss of income and unmet needs. People affected by cancer also shared worries in relation to job security and the disruption that this causes to daily functioning and broader family-related needs  Feelings of social isolation extend further beyond support within their home, to simple daily living needs or support when attending hospitals for cancer treatment.  Participants with endocrine-based cancers reported isolation negatively affected their quality of life [26] and others shared feelings of loneliness due to lack of social interaction during treatment |

**Appendix 13**: Summary of results on other aspects of cancer care during Covid-19 pandemic

| Adham M et al. 2022 | Head and neck cancer | Preoperative screening needed for patients undergoing surgery during the pandemic, with different number of PCR negative tests required before surgery in head and neck cancer. Some centers also used antibodies test. |
| --- | --- | --- |
| AlkatouI et al. 2021 | Any cancer site, breast cancer, colorectal cancer, gynecological cancer, lung cancer, colon cancer | The effect of delayed cancer diagnosis will not be perceived in the immediate future alone; premature deaths may occur as long as ﬁve years later. Delayed cancer screening is estimated to cause the following additional numbers of cancer deaths secondary to breast, esophageal, lung, and colorectal cancer, respectively: 54,112–65,756, 31,556–32,644, 86,214–95,195, and 143,081–155,238 in the worldwide |
| Alom S et al., 2021 | Any cancer type, and per different types | 6 core themes that encompassed common cancer service  intervention adopted by institutions were identified: (1) Testing and Tracking, (2) Outreach and Communication, (3) Protection, (4) Social Distancing (5) Treatment Management, (6) Service Restructuring. Many institutions have adopted various strategies to safeguard their patients and staff and streamline service provision, however the extent of success of these interventions is still unknown. |
| Cosimo SD et al. 2022 | Any cancer site, and cancer-specific including brain, head and neck, gynecological,  breast, hepato-bilio-pancreatic, hematological, colorectal, skin,  pediatric, urinary tract, esophagogastric, neuroendocrine, lung  and soft tissues cancers. | - Virtual visits were implemented by the majority (72%) of center - Routine use of PPE by patient and healthcare personnel was reported by 81% and 80% of centers, respectively; systematic SARS-CoV-2 screening by nasopharyngeal swabs was reported by only 41% of centers. |
| Donkor et al. 2021 | Any cancer type | There were four themes that emerged: preparing and equipping  staff; reinforcing infection prevention and control policies; strengthening coordination and communication; and maintaining physical distancing  Studies reported that radiotherapy centres had formed COVID-19 response multidisciplinary team; maximized the use of telehealth; adjusted the layout of waiting areas; divided staff into teams; dedicated a room for isolating suspected cases; and adopted triage systems. |
| Gadsden T et al. 2022 | Any cancer site and cancer-specific, including cervical cancer (n=1), paediatric (n=1), oral (n=1), blood (n=1), gastrointestinal (n=1) and head and neck (n=1) cancers | Of the 17 studies, only two reported on the impact of mitigation measures to maintain service provision during the pandemic. In the absence of any national guidelines in India, Mallick and colleagues prioritised radiotherapy treatment for oncology patients, continued services for patients already undergoing treatment and deferred new starts for adjuvant therapy. Additionally, a staff rotation policy was implemented to ensure that human resources could be redeployed to prevent delays and deliver full services for those with the highest priority. Although outpatient consultations dropped by 58% during lockdown, more than 90% of high-priority cancer treatments (specifically radiotherapy and chemotherapy) were implemented as planned. |
| Gascon L et al. 2020 | Head and Neck cancer | Recommendations include adjustments regarding new patients’ referral such as performing a pre-appointment triage and working in telemedicine when possible, testing Covid-19 status prior to surgery, use of personal protection equipment, as well as presence of only essential staff in the operating room and limiting the number of patients in the hospital. |
| Hojaij FC et al. 2020 | Head and Neck, and otorhinolaryngology | The use of adequate PPE (N95 mask or PAPR) with complete gown in aerosol-producing procedures should become mandatory until there is control of the epidemy, that is, vaccine or effective antiviral drugs. Endoscopic exams should only be performed if their result may change the patient’s treatment; otherwise, patient’s evaluation should be restricted to clinical exam. Evaluation of the COVID-19 status of all inpatients, especially surgery patients, should be performed to access transmission risk. Telemedicine can help reduce hospital and outpatient visits. |
| Majeed A et al. 2022 | Pediatric cancer | The reorganization of hospital services was reported to have  mixed results. In several instances, hospitals were able to restructure services  to better provide care for patients, including guidelines for social distancing. Pediatric cancer. In a tertiary government healthcare center in India, conversion to telemedicine greatly benefitted their patients; in the period between April-July 2020, teleconsultations rose exponentially (32 in April to 197 in July, 2020). Chemotherapy plans and prescriptions issues were also managed through email and were especially helpful for patients who lived far from the medical center |
| Moujaess E et al. 2020 | Any cancer site | Telemedicine was encouraged. Measures to protect medical staff are proposed because this indirectly impacts patients’ safety. These measures consist of prioritizing laparoscopic procedures in cancer surgery to minimize the exposure to aerosolized specimen and limiting endoscopic diagnostic procedures to the necessary with application of strict protective measures particularly in bronchoscopy. Some medical and imaging oncology wards were completely re-organized to safely accommodate cancer patients. A detailed description of Chinese, Italian and French experience is provided, as well a summary of international guidelines for management and care of cancer patients. |
| Pacheco RF et al. 2021 | Any cancer site, including breast cancer, head and neck cancer and lung cancer. | - Five multiple strategies and four single strategies were reported, and the possible effects of mitigating delays and disruptions in cancer care because of COVID-19 are inconsistent. - The findings emphasize the infrequency of measuring and reporting mitigation strategies that specifically address patients’ outcomes and thus a scarcity of high-quality evidence to inform program development. |
| Rohilla KK et al. 2021 | Any cancer site | - During a pandemic, the government of India started telemedicine consultations in which there were no social contacts, which is a valuable asset for both patients and healthcare professionals. The risk-adapted model was selected for it, i.e., patients who were already registered in the hospital and who were on follow-up can easily assess telemedicine consultation in the near future before the pandemic is over. |
| Lignou S et al. 2022 | Pediatric cancer | - A study on the impact of delays in cancer diagnosis in adults and children estimated that between 3291 and 3621 avoidable deaths will have occurred from 5 cancer types in the 5 years after diagnosis compared with the pre-pandemic period. An additional 59,204–^2 6-12 16-30^years of life lost will be attributable to delays in cancer diagnosis alone as a result of the first COVID-19 lockdown in the UK |
| Pararas N et al. 2022 | Colorectal cancer | - The number of patients presenting with metastases during the pandemic was significantly increased (OR 1.65, 95% CI 1.02–2.67, p = 0.04), with no differences regarding the extent of the primary tumor (T) and nodal (N) status. Patients were more likely to have undergone neoadjuvant therapy (OR 1.22, 95% CI 1.09–1.37, p < 0.001), while emergency presentations (OR 1.74, 95% CI 1.07–2.84, p = 0.03) and palliative surgeries (OR 1.95, 95% CI 1.13–3.36, p = 0.02) were more frequent during the pandemic. There was no significant difference recorded in terms of postoperative morbidity. |
| Tang G et al. 2022 | Colorectal cancer | - Covid-19 pandemic has not led to a deterioration in the surgical outcomes of colorectal cancer surgery or reduction in the quality of cancer removal. There was no statistically significant difference [OR, 0.90; 95% confidence interval (CI), 0.80, 1.01; p = 0.07] in the overall incidence of postoperative complications between patients in the COVID-19 pandemic group and those in the pre-COVID-19 pandemic group, with low heterogeneity between studies (I2 = 26%, p = 0.22). Meta analysis of the four studies showed no significant difference with regard to conversion rate. The result was OR = 1.07; 95% CI, 0.76, 1.52; p = 0.70 with high heterogeneity (I2 = 31%). Data on the anastomotic leakage rate were described in five studies. When colorectal cancer surgery was performed during the COVID-19 pandemic, this did not increase the incidence of anastomotic leakage (OR, 0.71; 95% CI, 0.43, 1.16; p = 0.17; I2 = 0%). The pooled effect sizes of the eight studies showed no significant difference in mortality (OR, 1.27; 95% CI, 0.92, 1.75; p = 0.14; I2 = 0%) between the two groups. Two studies reported on ICU demand rate. There was no significant difference in the ICU demand rate (OR, 0.73; 95% CI, 0.29, 1.85; p = 0.51; I2 = 0%) between the two groups. Four studies described R1 resection rate. There were no significant differences in the R1 resection rate (OR, 0.46; 95% CI, 0.11, 1.90; p = 0.28; I2 = 0%) for colorectal cancer surgery performed during the COVID-19 pandemic compared with that pre-pandemic. A meta-analysis of five studies did not show any significant differences in mean lymph node yield (MD, 0.16; 95% CI, −2.26, 2.59; p = 0.90; I2 = 54%). Colorectal cancer surgery during the COVID-19 pandemic did not increase the length of hospital stay (MD, −0.05; 95% CI, −2.28, 2.19; p < 0.00001; I2 = 98%) compared with that before the pandemic. The results of the sensitivity analysis showed that no single study significantly affected the overall effect size for postoperative mortality, conversion rate, mortality, ICU demand rate, R1 resection rate, anastomotic leakage rate, mean lymph node yield, and length of hospital stay. The total effect size for postoperative mortality changed (OR, 0.88; 95% CI, 0.78, 0.99; p = 0.03; I2 = 0%) when the study by Uyan et al. was excluded |
| Thomson JD et al. 2020 | Hypofractionated radiation therapy for any cancer sit, and cancer-specific including breast, central nervous system, cutanous, nonmelanoma, upper gastrointestinal, lung, lower gastrointestinal, genitourinary, gynecology, head and neck, hematologic, lymphoma, pediatrics, general palliative and sarcoma | - A large number of publications recommended hypofractionated radiation therapy schedules across numerous major disease sites during the COVID-19 pandemic, which were supported by a lower quality of evidence than the highest-quality routinely used dose fractionation schedules. For site-specific curative and site-specific palliative schedules, there was a significant shift from established higher-quality evidence to lower-quality evidence and expert opinions for the recommended schedules (P Z .022 and P < .001, respectively). For curative-intent schedules, the distribution of quality scores was essentially reversed (highest levels of evidence "pre-COVID" vs "in-COVID": high quality, 51.4% vs 4.8%; expert opinion, 5.6% vs 49.3%), although there was variation in the magnitude of shifts between disease sites and among specific indications |
